# Rapid realist review of virtual wards for people with frailty

**DOI:** 10.1101/2023.04.18.23288729

**Authors:** Maggie Westby, Sharea Ijaz, Jelena Savović, Hugh McLeod, Sarah Dawson, Tomas Welsh, Hein Le Roux, Nicola Walsh, Natasha Bradley

**Affiliations:** The National Institute for Health Research, Applied Research Collaboration West (NIHR ARC West), University Hospitals Bristol NHS Foundation Trust, Bristol BS1 2NT, UK; Bristol Medical School, University of Bristol, Bristol BS8 2PS, UK; RICE – The Research Institute for the Care of Older People, Bath; Royal United Hospitals Bath NHS Foundation Trust; Churchdown Surgery, Parton Rd, Churchdown, Gloucester GL3 2JH; NHS England and NHS Improvement South West; One Gloucestershire Integrated Care System Quality Improvement; Centre for Health & Clinical Research, University of the West of England, Bristol BS16 1DD, UK

## Abstract

**Background:** Virtual wards (VWs) deliver multidisciplinary care at home to people with frailty at high risk of a crisis or in-crisis, aiming to mitigate the risk of hospital admission. Different VWs models exist and evidence of effectiveness is inconsistent.

**Aim:** We conducted a rapid realist review to identify different types of VWs, and to develop explanations for how and why VWs could deliver effective frailty management.

**Methods:** We searched published and grey literature to identify evidence on VWs for frailty, based in Great Britain and Ireland. Information on how and why virtual wards might ‘work’ was extracted and synthesised in two rounds with input from clinicians and patient/public contributors, generating 12 hypothesised context-mechanism-outcome configurations.

**Results:** We included 17 published and 11 grey literature documents. VWs could be short-term and acute (1-21 days), or longer-term and preventative (3-7 months).

Effective VW operation requires common standards agreements, information sharing processes, an appropriate multidisciplinary team that plans patient care remotely, and good co-ordination. VWs may enable delivery of frailty interventions through appropriate selection of patients, comprehensive assessment including medication review, integrated case management, and proactive care. Important components for patients and caregivers are their communication with the VW, their experience of care at home, and feeling included, safe and empowered to manage their condition.

**Conclusions:** Insights gained from this review could inform implementation or evaluation of VWs for frailty. A combination of acute and longer-term VWs may be needed, within a whole system approach. An emphasis on proactive care is recommended.

## INTRODUCTION

People with frailty are at risk of unpredictable deterioration in health, and minor stressor events can lead to crises, care dependency and hospital admission. [1, 2] The UK has an ageing population, with an increasing prevalence of frailty, [2, 3] and there is recognition of the need for innovation in frailty management. [1, 4]

People with frailty form a diverse group, with different types and levels of need and support, for both health and social care. They may depend on others for activities of daily living, and both hospital admission and delayed discharge can occur because of a lack of community services. [5]

Delivering services to manage frailty requires a multidisciplinary team (MDT) that can provide a tailored, whole-person approach to diagnosis, assessment and treatment, aiming to stabilise frailty, intervene with crises, and prevent exacerbations. [1, 2, 6-8]

The virtual ward (VW) model of care combines components of care under a common scheme and delivers multidisciplinary preventative care to patients in their own homes, aiming to mitigate their risk of unplanned hospitalisation. In the UK, longer term (several months) VWs for frailty or long-term conditions have been introduced since the 2000s. [9-12] The term, ‘virtual ward’ is, however, used to cover a variety of models.

Recently, building on VWs for Covid-19, [13] NHS England has issued guidance on short-stay (a few days) VWs for patients with ‘acute exacerbations of conditions related to frailty’, with planned roll-out of VWs for frailty. [14]

Evidence of VW effectiveness is limited. Five studies [10, 15-18] and one systematic review [19] that compare VWs with usual care (one UK-based [10]) report inconsistent findings. Suggested explanations for poor effectiveness include failure in the functioning of the MDT, [10] indicating there may be crucial mechanisms by which VWs ‘work’ to improve patient outcomes.

### Rationale

In view of the complexity of VWs and the variability in quantitative findings from different countries, we were interested in how and why VWs could improve the management of frailty. This review seeks to explain how the context of the patient and the VW shape the outcomes of the intervention. Building this knowledge may help inform the successful implementation and evaluation of VWs for frailty.

Rapid realist reviews are suitable for investigating a defined topic area for a clear purpose, such as informing policy, and integrating different types of evidence including quantitative and qualitative, and published and grey literature. [20] Focussing primarily on the UK, this project planned to synthesise relevant evidence, producing a set of initial programme theories explaining ‘what works, for whom, and in what circumstances?’ [21] We aimed to describe the different models of multidisciplinary VWs in operation in the UK, and to investigate mechanistic factors that impact effective VWs in different contexts.

## METHODS

Full details are reported in Appendix I. [20, 22]

Preliminary scoping of the literature informed development of the review protocol, including our definition of VWs.

### Inclusion criteria

VWs were defined as (Appendix II): [12, 19, 23]

- The VW provides care to patients in their own homes in the community,
- A multidisciplinary team makes decisions/plans care remotely from the patient,
- The multidisciplinary team provides oversight of patient care.

Other inclusion criteria were: (i) people with frailty, multi-morbidities or older adults; (ii) set in Great Britain and Ireland; and (iii) evidence suitable for theory building.

We excluded VWs in care homes, children, people with Covid-19 or a single condition (e.g., cystic fibrosis).

### Searching and selection of documents

We conducted an initial Ovid multi-file search of the main medical/healthcare databases (MEDLINE, Embase, PsycINFO) to identify published academic literature (up to 8 November 2021) (Appendix III). We also searched for grey literature. Post-stakeholder consultation, we updated and extended the search and conducted forward citation searches (24 June 2022). Initial search terms related to multidisciplinary, virtual wards, and frailty/older people; later, hospital-at-home terms were added. Four authors screened titles and abstracts and full text, identifying relevant core documents, rich in information.

### Data extraction and synthesis

Three authors extracted data from the core documents, generating ‘if-then-because’ statements that captured relevant causal insights. These statements were organised into 21 topic areas, corresponding to different components of the VW or relevant contextual aspects.

Preliminary ‘context-mechanism-outcome configurations’ (CMOCs; Box 1) were developed from if-then-because statements, and then elaborated and refined using all the evidence, alongside further stakeholder engagement (Appendix IV).

#### BOX 1

Definitions

- **Context**: backdrop of the intervention and variations of this across sites, which existed before the VW implementation and are outside of the mandate of service redesign (e.g., policy, staff skills, IT systems).
- **Mechanism:** reasoning of stakeholders in response to resources offered by the intervention (e.g., trust and motivation to act).
- **Outcome:** includes intended and unintended outcomes of interest, such as: hospital admissions, safety, clinical outcomes, resource use, patient and caregiver satisfaction, etc.
- **CMO Configurations (CMOCs):** propositions explaining how the interaction between contexts and mechanisms can lead to outcomes of the intervention (i.e., VWs for frailty).

### Stakeholder engagement

Stakeholders were engaged in two stages. Initially, we presented a diagram of the patient pathway (Figure 1) to one clinician and two public contributors, requesting feedback, and facilitated discussion on the topic areas of the if-then-because statements. This generated new statements.

**Figure 1:**
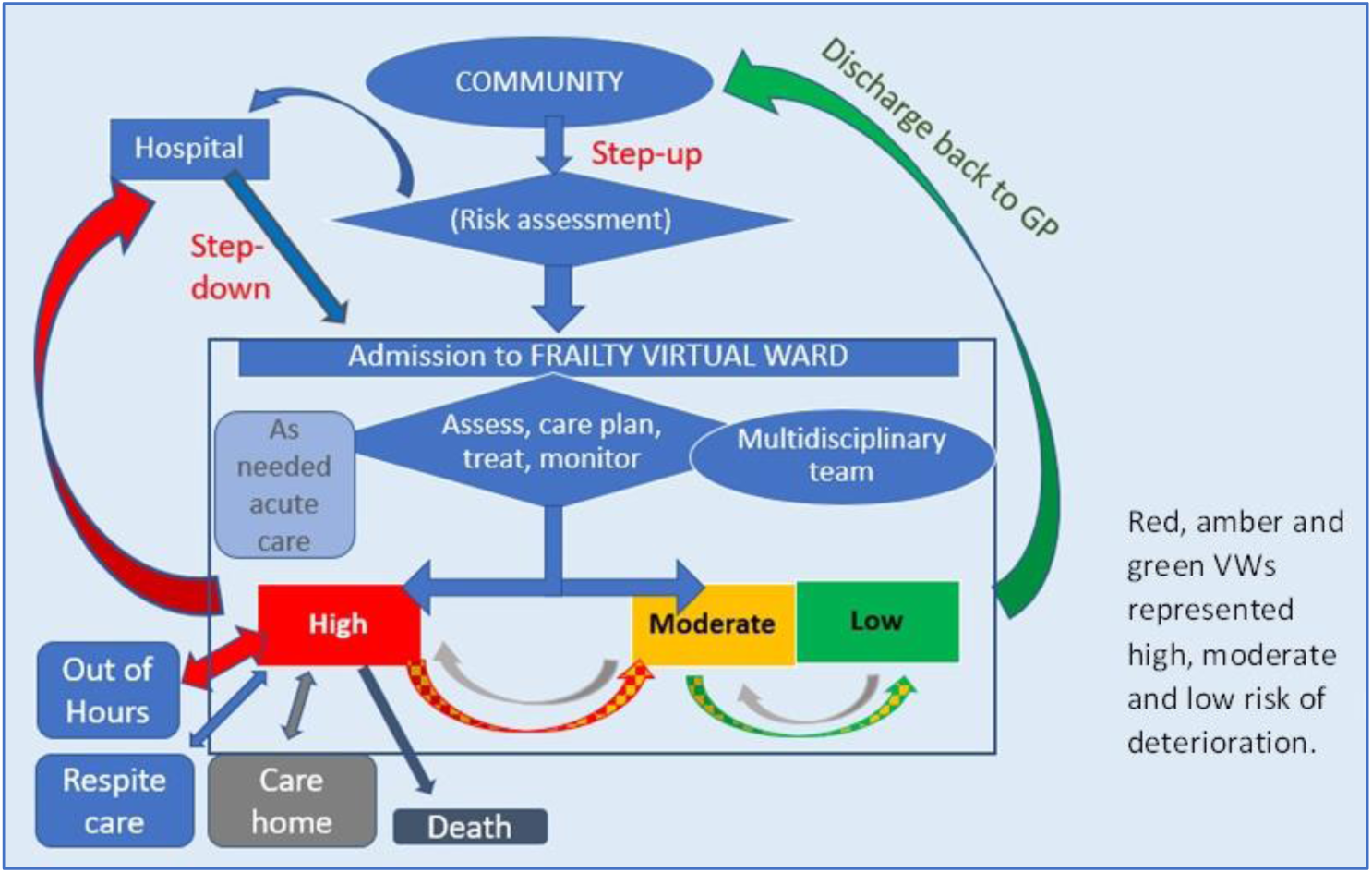
Overview of the VW model.

Stage two involved three clinicians with experience of VWs and five public contributors with experience of frailty. We presented draft CMOCs and made comparisons with NHS England guidance. [14] Subsequently, we broadened the review to acute VWs.

### Changes to the review process

We initially excluded acute ‘hospital-at-home’ interventions, but, following stakeholder feedback, added acute interventions if they met our VW definition. Resource constraints prevented us from performing detailed rigour assessment. We initially proposed a large stakeholder consultation, but adapted it to small online groups because of resource and scheduling limitations and Covid-19 precautions.

We had intended to include only UK-based VW models, but extended this to seminal work from Dublin by Lewis et al that had influenced VW development in the UK.

## FINDINGS

We describe document characteristics, types of VW, and summarise 12 CMOCs under three main themes. Full details of the CMOCs are in Appendix V.

### Document characteristics

The search process is depicted in Figure 2. Details of included documents are in Appendix VI. We included eight core documents in stage 1 [11, 12, 24-29] and 20 further documents in stage 2: nine published [10, 30-37] and 11 grey literature. [23, 38-47]

**Figure 2:**
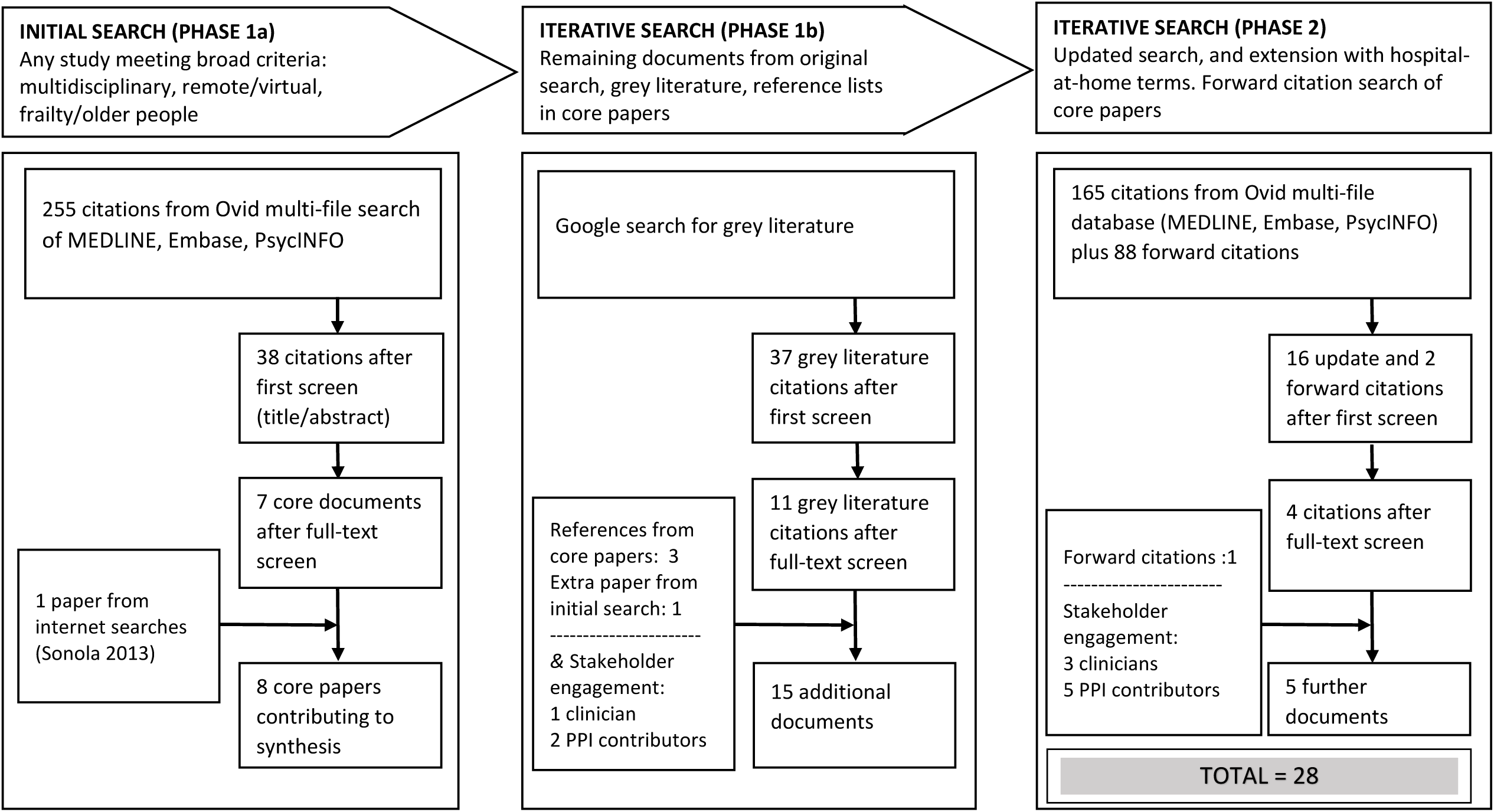
Flow diagram.

Fifteen documents described longer-term VWs, [10-12, 23-26, 28, 29, 31, 32, 34, 36, 44, 45] ten short-term VWs, [27, 30, 33, 35, 37, 38, 40, 42, 43, 47] two both models, [39, 46] and one was unclear. [41] Thirteen documents specifically included people with frailty. [11, 23, 28-30, 34, 36, 38, 42-44, 46, 47] Most studies are over five years old, four are from 2020-2022. [23, 27, 38, 43]

### Types of VW for frailty

VWs provide care at home for people acutely unwell with frailty or chronic conditions, at high risk of hospital admission. We distinguished two main types of VW: longer-term (3-7 months) and short-term (1-21 days) (Table 1).

**Table 1:**
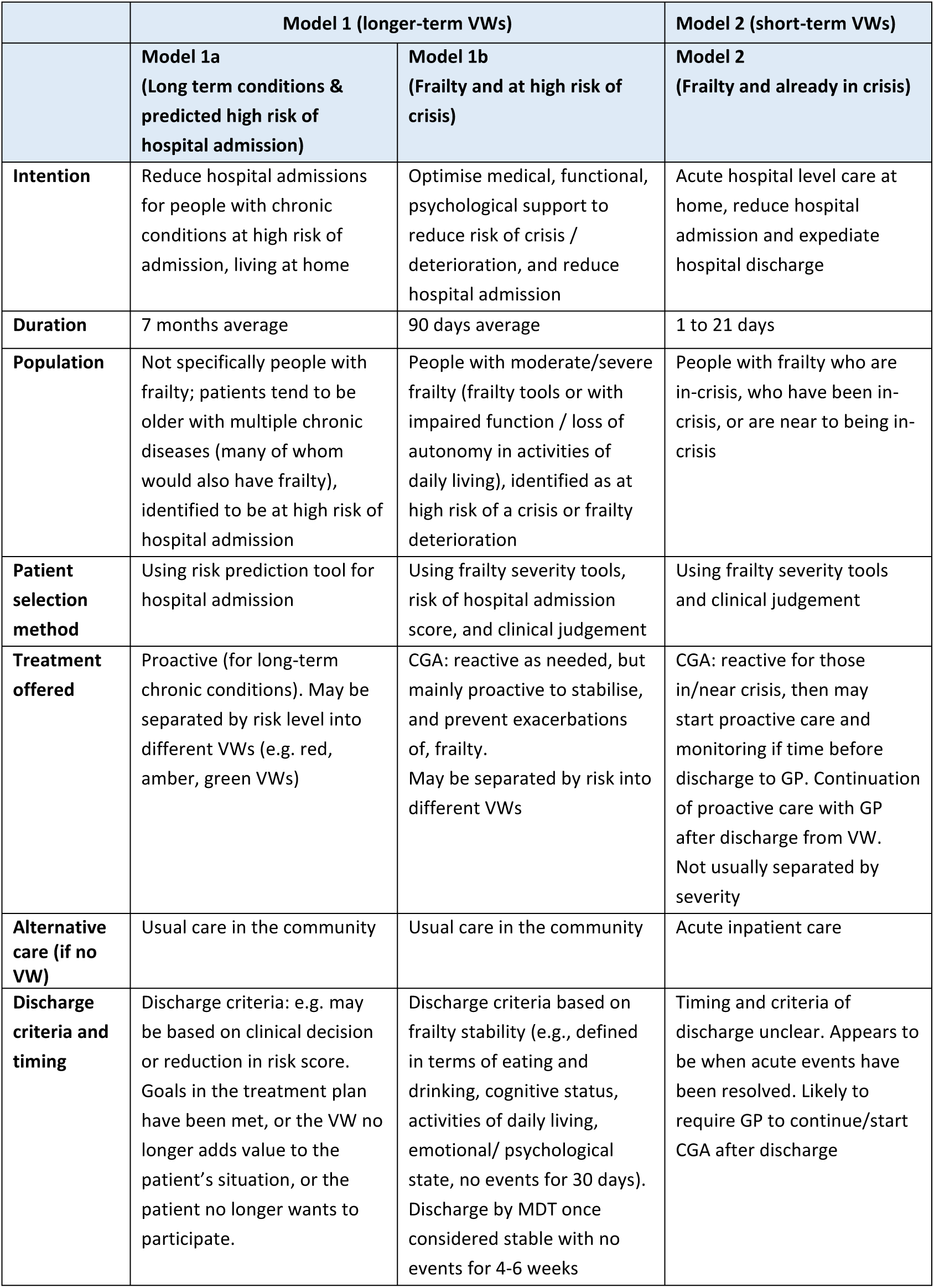
Types of Virtual Ward.

Originally, VWs (e.g. [12], model 1a) were intended to reduce hospital admissions by proactively treating older people with chronic conditions at high risk of admission. Patient selection was usually based on risk prediction modelling.

Later VWs focused on frailty (e.g. [11]; model 1b) and were intended to assess and stabilise frailty in people at high risk of a crisis. Reactive care was first offered to alleviate any acute frailty exacerbations, before focussing on proactive care to reduce risk of future crises, for example, the Comprehensive Geriatric Assessment (CGA). [48]

Longer-term VWs are alternatives to usual care in the community (for people at high risk of exacerbations). In some VWs a traffic light system (red/amber/green) is used to prioritise assessment, monitoring and intervention according to risk of deterioration.

Short-term VWs admit people with frailty already in-crisis or very near to a crisis (e.g. [30], model 2), offering acute reactive care. Proactive care may be started if time, but such VWs should plan for continuity after discharge to primary care. Short-term VWs are alternatives to inpatient hospital treatment. NHS England guidance on frailty VWs is based on this model. [14]

### CMOC Theme 1: VW Building Blocks

#### CMOC1: Common Standards Agreements

When there is sufficient motivation towards establishing a VW, common standards agreements can be developed between the different providers and specialities, such that legal and regulatory requirements of the different authorities can be met. (Context).

Common standards agreements cover topics such as patient eligibility, assessment procedures, care documentation, data protection, safeguarding, and discharge. Provided they suit the working practices and cultures of the different teams, having transparent agreements in place about the purpose and processes of the VW offers clarity on role expectations, encouraging confidence that the VW will function and not put patients at risk. (Mechanism).

Common standards agreements formalise collaboration because operational agreement underscores the communication and teamwork between professionals. This facilitates effective decision-making and case management, leading to improved efficiency. (Outcome).

#### CMOC2: Information Sharing Processes

Where there is enough progress towards IT integration and trust between providers, information sharing and ‘real-time’ data management processes can be established between the different organisations (including those external to the VW – out-of-hours and emergency services). (Context).

Information sharing processes can equip professionals with an accurate ‘whole system’ view of the patient record, and can increase confidence through having accurate information when needed. Patient and caregivers appreciate not repeating themselves; and may feel reassured that their clinical team is well-informed. (Mechanism).

Decisions can be better informed and timelier, which improves patient management because processes of care and access to appropriate interventions can be streamlined. (Outcome).

#### CMOC3: Multidisciplinary Team (MDT) composition and coordination

The expertise required for frailty management may be disparate across multiple teams, including primary care, community care, and speciality frailty clinicians. Key recognisable professionals can ‘champion’ working together in the VW. (Context).

MDT composition encourages professionals to trust that the model can provide safe and personalised care for patients at home. VW co-ordinators may facilitate teamwork, organise task sharing and liaise with both patient/caregivers and external organisations. Effective team composition and coordination means that all parties feel secure in the VW model. Professionals are willing and able to participate in the team, taking a shared approach to tasks and problem-solving. (Mechanism).

Patient management benefits from expertise and skills from different specialities and organisations. Team composition and coordination improves patient access to a range of interventions. Unnecessary or duplicated effort in patient care is reduced. (Outcome).

#### CMOC4: MDT meetings

The aims of the VW model and its implementation can motivate different teams and disciplines to work together. Regular MDT meetings, well-attended by team members facilitate MDT function, provided the professionals involved have sufficient capacity in their workload. (Context).

The MDT meets to discuss patients and plan their care, usually via technology or sometimes in-person. Meetings provide a forum for communication between specialist clinicians and the care teams providing hands-on care. Ideally VW professionals perceive these meetings to be worthwhile and participate in collaborative problem-solving. (Mechanism).

Collaboration improves holistic patient management by enhancing the effectiveness and efficiency of decision-making across different disciplines and providers. Supportive communication and task-sharing may provide learning and upskilling opportunities for staff. (Outcome).

#### Implications for VW planning

Sufficient motivation and co-operation between the teams involved is needed to develop and successfully introduce common standards agreements. These may need review and revision as the VW becomes more established over time.

Similarly, introducing effective IT integration requires perseverance and collaboration between organisations. Ineffective information sharing can mean duplication of effort and ‘silo working’ – frustrating for both staff and patients - and may impede the timeliness or appropriateness of decision-making.

Team composition varies according to the aims of the particular VW model and local patient need. It may include: geriatricians, physiotherapists, pharmacists, social workers, mental health professionals, voluntary sector, community organisations, and other clinical specialities (e.g. cardiology). The MDT usually meets remote from the patient, with decisions enacted by community teams. Successful communication and care documentation are essential. Ideally, all professionals feel confident they will have accurate information when they need it, facilitating prompt and well-informed decisions on patient management.

The VW facilitates shared learning across traditional role boundaries, enhancing collective capacity for patient care. Co-location of VW team members could increase their connectedness and joint working. However, poor understanding of VW aims could lead to role protectionism that undermines MDT functioning.

Effective MDT meetings are crucial for the VW to function as a forum for the integration and prioritisation of patient care. Online meetings can facilitate attendance and save time, but professionals involved must have time and capacity to attend. Disparity in attendance could delay decision-making and demotivate attendees.

### Theme 2: VW delivering the patient pathway

#### CMOC5: Patient selection

Against a backdrop of scarce resources, VWs should select and prioritise appropriate patients. GPs use frailty risk tools as part of the GP contract, and can identify people in the community who are in-crisis or who are nearing a ‘tipping point’ into crisis. (Context).

Selection into the VW may be informed by both clinician judgement and frailty risk tools or hospital risk prediction tools, which prioritise patients to the VW and provide a coherent rationale for their selection. Professionals perceive they can make a difference by working together to safely keep these patients at home. (Mechanism).

Selected patients receive timely and targeted management that could stabilise their condition or prevent a crisis, lowering risk of unplanned hospitalisation and reducing length of stay if admitted. (Outcome).

#### CMOC6: Comprehensive assessment and evaluation

The multidimensional needs of frailty require a holistic approach, such as the CGA. [48] MDT composition and functioning facilitate access to the interventions, specialists, and services required to be responsive to multidimensional frailty needs. (Context).

Patients receive a holistic ‘assessment’ -usually face-to-face with the VW co-ordinator – including use of appropriate screening tools and goal setting. The co-ordinator and MDT then prepare and enact a personalised management plan. The MDT feels confident in information from the assessment. (Mechanism).

Patient needs are identified, appropriate interventions are mobilised to meet them, and the patient receives timely access to specialists and healthcare services. Reduced duplication of effort compared with ‘siloed’ management may expedite access to interventions and could improve overall efficiency. (Outcome).

#### CMOC7: Medication management

Polypharmacy is common in people with frailty, and specialist input for medication management may be required for complete and effective medication reviews. (Context).

A personalised medication review at home, through the VW, enables accurate reconciliation of prescribed and non-prescribed medications, and provides opportunity to identify sensory impairment or side effects, explain any changes, and respond to concerns. Extra time to discuss medication management could improve patient and caregivers’ understanding of their treatment. (Mechanism).

Unnecessary polypharmacy can be identified and resolved safely. This could improve treatment adherence, lower the risk of adverse events, and reduce the treatment burden for some patients. (Outcome).

#### CMOC8: Intensive case management

People with frailty have complex health and social care needs. The VW brings together an effective ‘team of teams’ that can deliver multidisciplinary input within the patient’s home. Initially, interventions may be acute reactive treatment for frailty crises, such as delirium or immobility. (Context)

In-person visits and monitoring provide the VW with accurate and timely information, and progress is reviewed regularly during MDT meetings. Patients may be stratified by risk (e.g., using red/amber/green ratings), which determines the frequency of monitoring and MDT review. The VW is well-informed and responsive to patient needs. Ideally, patients/caregivers feel visible to the healthcare system in a way that feels safely supported. (Mechanism).

The VW can respond rapidly to changing patient needs and timely intervention is enacted. Monitoring and review mean that the VW can determine when a patient is stable and ready for discharge back to their GP. (Outcome).

#### CMOC9: Proactive care

Frailty is characterised by fluctuations in health, which can lead to frailty crises. Some VWs deliver proactive measures to people at high risk of a crisis, to prevent future deterioration or need for acute care. (Context).

Patients and caregivers receive proactive input such as: support for hydration, nutrition, and personal care; self-management strategies; advanced care planning; and as appropriate, mental health; falls prevention; physiotherapy and social support. VW professionals feel impactful in addressing potential issues, thereby preventing future crises. Patients and caregivers feel supported and ideally more confident in managing at home. Patients can be empowered to become active in their own care. (Mechanism).

Proactive care aims to stabilise frailty, and support patients and caregivers in planning for living with frailty in the longer term. Proactive care could prevent or mitigate the impact of future deterioration and crises, helping the patient avoid hospital admission, and potentially improving quality of life and patient safety. (Outcome).

#### Implications for delivering the patient pathway

Patient selection processes should be coherent with the aims of the VW and its common standards agreements. Professionals’ perceptions that the VW is prioritising the ‘right’ patients - taking an acceptable stance on risk of harm and likelihood of benefit - may be important for their trust and motivation in the VW. Conversely, the benefits of working in the VW may become less clear if patient selection is ineffective.

The VW co-ordinator involves the patient and/or caregiver in drafting the management plan, and works with the wider team to refine and deliver it. The VW can be a supportive learning environment that facilitates this way of working – however, this could be threatened if key team members cannot maintain regular communication necessary for integrated case management.

VWs stabilise frailty by facilitating timely, proactive interventions to mitigate future risk, and also provide as-needed acute care for crises. Some VWs may have insufficient capacity or time to stabilise frailty and rely on GPs to continue the treatment plan, which requires mechanisms ensuring good continuity of care at discharge from the VW.

### Theme 3: Patient and caregiver experience

#### CMOC10: Improved communication

VW processes enable effective communication and information sharing with the patient and/or caregiver, and provide a route to make contact out of usual working hours. Although most VWs do not provide 24-hour cover, alert systems notify the VW if patients attend emergency care/out-of-hours services. (Context).

The time to seek and receive assistance can be reduced through enhanced contact mechanisms. Consistent access to a known staff member (the VW co-ordinator) is reassuring for the patient and/or caregiver. The VW team can remain well-informed, which facilitates responsive treatment. (Mechanism).

Improved communication could boost patient and caregiver satisfaction and confidence in managing at home. Anxiety may be reduced through increased awareness of the support in place. (Outcome).

#### CMOC11: At home instead of hospital

Frailty-related crises may be alleviated by intervention, however extended or repeated stays in hospital could have negative consequences for the health and wellbeing of a person with frailty, and potentially their family or caregiver(s). Usually, patients and caregivers who feel comfortable and secure at home prefer to avoid being in hospital. (Context).

The VW facilitates integrated case management for people with frailty, so that appropriate and timely interventions can be delivered at home. Patients and caregivers feel supported and safe. (Mechanism).

Remaining in a familiar environment could enable patients and caregivers to maintain existing routines, such as for physical activity and social support. The disruption of hospitalisation is avoided which may contribute positively to health/wellbeing. (Outcome).

#### CMOC12: Caregiver experience

Patients and their caregivers have practical, informational and emotional support needs and may find it challenging to navigate complicated healthcare systems. (Context).

Where appropriate, caregivers are included in VW communication and shared decision-making, giving the VW more insight on the patient’s situation and on patient and caregiver needs. The caregiver feels supported by the VW and valued and listened to. (Mechanism).

Involvement in proactive care planning increases caregiver confidence in continuing to manage after patient discharge from the VW, and caregiver burden and stress is reduced. (Outcome).

#### Implications for patient and caregivers’ experience

VWs should aim to include the patient and/or their caregiver in decision-making without over-burdening them. Improved communication between the patient/caregiver and the VW, via a known point of contact (e.g. a well-informed, reliable co-ordinator), is expected to be reassuring. It is important to have clear communication on discharge and its timing.

Caregivers and patients ideally feel more confident because of VW intervention. However, revoking VW support at discharge may result in an increase in anxiety, especially if the patient or caregiver does not feel well-equipped by the VW to continue at home or proactive care is not established. Ideally, patients feel empowered to manage, but conversely, if VW input means patients feel less enabled, they could lose confidence, potentially increasing stress for both patients and caregivers.

Effective continuity of care with primary care is important at discharge for the patient/caregivers to regain confidence living outside the VW. Communication with the GP should support continuation of the management plan. Where this is missing, patients and caregivers may be left with uncertainty and heightened anxiety.

In some cases, the home environment may not be safe, and hospital may be more suitable. It may be that caregivers are unable to take on additional responsibilities, for example, for patients experiencing delirium or other frailty crises, or the home setting is unsafe for the delivery of acute interventions.

## DISCUSSION

This rapid realist review drew from 28 documents and the experiences of clinicians and public contributors. We generated evidence-based theories: first of how various VW components and patient and caregiver involvement contribute to effective operation of VWs, and second, how these components combine as one entity to deliver frailty interventions.

In a field in which there is uncertainty around what constitutes a VW, we refined, with stakeholders, a definition of VWs as a model of service delivery in which an MDT meets and plans patient care remote from the patient, and co-ordinates multidimensional interventions at home for people acutely unwell with frailty (at high risk of crisis or in-crisis).

### Summary of findings

We identified two types of VW, which differ in their aims, duration and the patients’ stability. Longer-term VWs (3-7 months) focus on proactive treatment to stabilise frailty and prevent a crisis in people at high risk of a crisis or of hospital admission. Short-term VWs (1-21 days) treat people with frailty already in-crisis instead of acute care in hospital. Primarily, they provide rapid access to acute reactive care, and, if time, start proactive care before discharging patients to GP care. Short-term VWs may be viewed as a service that catches people who fall through frailty management gaps. With increasing prevalence of frailty, short-term VWs may not be sustainable.

Fundamental to VW functioning are key building blocks with their underlying mechanisms. These comprise: robust information sharing and common standards agreements that the teams can understand and work within; multidisciplinary teamwork, featuring remote MDT decision-making meetings alongside in-person care; and effective co-ordination, with links to external services (such as out-of-hours). Also important are good relationships within the VW, in-person contact, and involvement and inclusion of patients and caregivers.

Pertinent mechanisms relate to the motivation of professionals to work together and their ability to do so. Ideally the VW operates as a ‘team-of-teams’ providing mutual support, trust in shared goals, and benefit from reciprocal learning. Perceptions of patient safety and benefit, starting small and taking time to introduce formal agreements and learn new ways of working may be necessary for professionals to ‘buy in’ to the VW model. Also essential is good communication between patients, caregivers and staff, and enabling them to feel safe at home and empowered to manage their own care.

Ideally, the VW components combine to ensure the VW as a whole can deliver interventions to people acutely unwell with frailty. Interventions include selection into the VW, comprehensive assessment and evaluation, integrated case management and proactive care.

VWs are not usually 24-hours. For some people with frailty (e.g., those with delirium), limited evidence suggests an impact of frailty symptoms on caregivers, who must take on a bigger role (particularly out-of-hours), and may feel unable to cope at home, leading to caregiver stress and burnout, and patient hospitalisation. Sometimes acute care in hospital is the best arrangement.

### Whole system context

VW delivery of frailty interventions should be considered in a whole system context, including transfer of care into and out of the VW.

In longer-term VWs, patients are mainly referred from primary care, following set criteria. In short-term VWs, referrals are likely urgent, and may be from primary care, emergency services or early discharge from hospital. Before reaching a crisis, patients with frailty may have been treated in the community to prevent deterioration, possibly in GP-managed ‘pre-wards’.

Timings and arrangements for discharge to GP care differ: in longer-term VWs, discharge is when the MDT determines patients to be stable following proactive care; the co-ordinator arranges good continuity of care. In short-term VWs, discharge may occur when acute events have been resolved; CGA may have been initiated in the VW, but there is insufficient time to establish proactive care. Effective continuity of care on discharge to primary care therefore becomes essential.

It may be that a combined approach to community care is helpful. One study reported such a model, comprising a longer-term VW, urgent care and a care home. [46]

### Cost implications

All VWs require investment of finances and time. This investment could be offset if VWs (compared to alternative care) reduce unplanned or prolonged hospital admission, duplication of effort between care providers, and/or improve the efficiency of decision-making. Different types of VWs would vary, particularly in terms of staff costs and length of stay in the VW. The ability of VWs to impactfully prevent or shorten episodes of hospitalisation may be highly contingent on resources being available elsewhere (e.g., domestic care workers). Resource demands should take a broad perspective, including out-of-pocket costs for the caregiver.

Current systems and services can be too reactive and hospital-centric, becoming unaffordable as the population changes. [39] Limited financial resources in both hospital and community care are the background of VW implementation in the NHS, [35] recognising that current care pathways are unsustainable and a more proactive system of care is required. [39] Improvements in frailty management could become cost-saving at the system level if people can be reached before a crisis and better supported to maintain their independence at home for longer.

### Comparison with other work

Existing systematic reviews of effectiveness of VWs are limited, [19] are restricted to RCTs (of which there are few for frailty VWs), and do not answer questions about how and why VWs are effective. In contrast, this review draws on a range of document types, including grey literature, to answer these questions. Our work may complement systematic reviews of RCTs of community-based complex interventions, which use techniques such as component network meta-analysis to determine which components are important. [49]

In December 2021, NHS England produced guidance to introduce ‘virtual wards’ for patients with ‘acute exacerbations of conditions related to frailty.’ [14] Recent work has also focused on short-term VWs or hospital-at-home models for acute care: a rapid evidence synthesis of systematic reviews of acute VWs, hospital-at-home and remote monitoring, across all countries, [50] and the British Geriatrics Society’s position paper on VWs for older people with frailty. [51] Our review included a broader range of types of VW and was not limited to the more topical short-term VWs.

This rapid realist review is the first to explore how, why and for whom VWs may deliver effective frailty interventions. The findings show similarities with that of a larger realist synthesis on inter-organisational healthcare, which reports that collaborative leadership ‘works’ when there is trust between the parties involved, faith in the proposed model of care, and confidence in its ways of working. [52]

### Strength and limitations of our work

The review explores the underlying mechanisms for VWs. We followed RAMESES standards and involved clinicians and patient/public stakeholders. However, we were unable to recruit patients with lived experience of a frailty VW, and perspectives on the caregiver experience were limited. The rapid realist review focussed on the UK, so was directly relevant to current NHS practice.

One limitation is that most evidence came from before the Covid-19 pandemic and/or from periods when the UK health system was different in structures and pressures. For example, the role and expectation of technology has changed rapidly, but was not captured in most included documents.

We did not formally assess rigour of included studies due to resource limitations. We consider its impact on our findings minimal as the synthesis generates hypotheses rather than evaluating effectiveness.

### Conclusions

This rapid realist review outlines different types of VW for people with frailty. We report 12 context-mechanism-outcome configurations that specify important VW components and the circumstances in which these lead to intended outcomes.

Preferably, people with frailty at high risk of crisis are identified in primary care before reaching a crisis, and receive proactive assessment, monitoring, and support to self-manage, thereby preventing crises. This care may be within a longer-term VW, or could be in GP-managed ‘pre-wards’ before admission to acute care VWs. Short-term VWs that admit people with frailty already in-crisis may be a safety net for people who fall through frailty management gaps. This may not be sustainable as the prevalence of frailty increases.

Our findings could inform future decisions regarding service planning, evaluation, and implementation of VWs for frailty. There is insufficient evidence on the sustainability of VW models, experiences of caregivers, or the impact of social inequalities, all of which should be examined further.

### Recommendations

Establishing and maintaining a VW should involve formal collaboration agreements and adoption of new ways of working. Time and resource are required and should be planned into professionals’ work schedules. Patient safety and benefit are important to professionals - this may necessitate ‘starting small’ so professionals can ‘buy-in’ to the VW model and adapt to change.

The risk of caregiver stress, anxiety, or burnout in some situations (e.g., delirium) should be considered, especially where the situation impacts disproportionately on caregiver responsibilities, including after hours when VWs may not provide support. For some patients, hospital with 24-hour care may be the best place.

A whole system approach to effective frailty management should be considered, with an emphasis on continuity of care including referral and discharge experiences. There may be a role for a combination of VW types within the system.

Frailty VWs should emphasise proactive care that can identify and reduce risk of future crises as part of a sustainable long-term view of frailty management. This, done in the context of rigorous evaluation, would have potential to improve quality of life for patients and their caregivers, alongside reducing the system burden of frailty care in the NHS.

## Data Availability

Data produced in the present work are contained in the manuscript or the supplementary appendices. Any additional data are available upon reasonable request to the authors

## Acknowledgements

We wish to acknowledge the helpful contributions from their experience and perspective, of the eight public contributors and one additional clinician, alongside support from our PPI coordinator in finding suitable contributors.

