## Supplementary Appendices for "Rapid realist review of virtual wards for people with frailty"

### APPENDIX I – Methods

This methods section follows the RAMESES publication standards. [1]

#### 1.1. Rationale for using a rapid realist review

Realist research methods are valuable in approaching complex interventions that have heterogenous or context-depending outcomes. Realist reviews ask “what works for whom, under what circumstances, how and why?” by conducting a theory-led review of diverse evidence, developing explanations of the interactions between context and mechanisms of an intervention in leading to outcomes. [2] Different types of evidence and research design may be incorporated, including peer-reviewed literature and grey literature such as service evaluations and blogs.

In the case of virtual wards (VW), their operation appeared to be underpinned by multiple components working together to deliver frailty management that is intended to improve outcomes for patients and the healthcare system. However, there is variability in VW models and results reported. The ways in which VW operation depends on context and the relevant mechanisms at play remained obscure and led to this review.

Rapid realist reviews are suitable to investigate a defined topic area for a clear purpose, such as informing policy. They are distinct from realist reviews because they are conducted with a programme-specific aim in mind, rather than with the goal to produce transferable/generalisable findings. [3] In this case, we focus on VWs for frailty in the UK. Our rapid realist review will contribute evidence-based explanations of the processes of setting up and delivering VWs for people with frailty.

#### 1.2. Process overview

After scoping the literature (NW and HM), writing the protocol (NW) and determining the review questions, we conducted two main rounds of literature searching and synthesis, and engaged with stakeholders at both stages.

At the first stage we worked with ‘if-then-because’ statements. After reviewing eight ‘core documents’ (MW, NB, SI, NW), project team brainstorming (MW, NB, SI, NW, HM, JS), meeting with one clinician (one meeting) and two public contributors (one meeting), we had 113 unique ‘if-then-because’ statements (partial or complete) and these were organised under 21 topic/component headings (NB).

During the second stage we developed ‘context-mechanism-outcome configurations’ (CMOCs), bringing together the if-then-because statements under each of 12 headings, informed by our definition of context, mechanism and outcome (Box 1) (NB). We then discussed these CMOCs with three clinicians (five meetings in total) and five public contributors (one meeting), and also added data extracted from the remaining identified evidence sources (MW, SI).

In the evidence, we looked in particular for new information that refuted or elaborated on the preliminary CMOCs. Through iteration and refinement this gave a final set of 12 CMOCs.

See Appendix III for the search strategy and Appendix IV for an overview of CMOC development, and Appendix V for detail of CMOCs.

BOX 1: Definitions

- **Context:** the backdrop of the intervention and variations of this across sites, which existed before the VW implementation and are outside of the mandate of service redesign (e.g., policy, staff skills, IT systems).
- **Mechanism:** the reasoning of stakeholders in response to resources offered by the intervention (e.g., trust and motivation to act). The protocol outlined nine potential stakeholder groups: medical consultants; GPs; physiotherapists; occupational therapists; community nurses; social care representatives; patients; carers; and service commissioners.
- **Outcomes** included intended and unintended outcomes of interest, including but not limited to such as: hospital admissions, safety, clinical outcomes, resource use, patient and caregiver satisfaction, costs, inclusivity, etc.
- **CMO Configurations (CMOCs):** propositions explaining how the interaction between contexts and mechanisms can lead to outcomes of the intervention (i.e., VWs for frailty).

#### 1.3. Changes to the review process

We initially considered hospital-at-home to be a distinct model of care to VWs. However, following stakeholder discussions and interrogation of NHS England guidance, we broadened the scope to include hospital-at-home for frailty, provided it met our inclusion criteria and definition of VWs (see Appendix II). This decision meant that we extended our search terms and updated the search. We had expected to refine our findings through a formal stakeholder consensus exercise involving 20-30 people. However, due to resource limitations of this project, precautions against the transmission of Covid-19, and the ongoing pressures in the healthcare system, we instead conducted small online groups and involved a smaller number of stakeholders than originally intended.

The protocol identified nine stakeholder groups which could be relevant for theorising (medical consultants; GPs; physiotherapists; occupational therapists; community nurses; social care; patients; carergivers; and service commissioners). There was insufficient evidence available to investigate mechanisms for each of these stakeholder groups.

#### 1.4. Further details

Literature scoping

Two authors (NW and HM) collated initial literature which was discussed with the entire team and further scoping literature was obtained through searching references and through contacts in the field. We defined the scope of the project, the research questions, and a search strategy based on this literature. NW wrote the protocol.

Search process

Literature searching was done in two stages in collaboration with an information specialist (SD).
In Stage 1a, we conducted an initial Ovid multi-file search of the main medical/healthcare databases (MEDLINE, Embase, PsycINFO) to identify published academic literature (all years to 8 November 2021), using terms relating to multidisciplinary teams, remote/virtual care, frailty/older people (see Appendix III for full search strategy).
In stage 1b, we examined the reference lists of other systematic and realist reviews (backward citations) and searched for grey literature, running iterative, informal searches on Google/Google

Scholar and followed related links.
In stage 2 – post-stakeholder consultation - we updated and expanded the search (24 June 2022) to include terms for hospital-at-home. We also conducted a forward citation search at this time, using Web of Science and Google Scholar to identify new research citing key documents identified previously.

Two authors screened the titles and abstracts identified in stage 1 for relevancy (NW, MW) and one author resolved any differences (SI). Then three authors assessed the full papers (NB, SI, MW) for inclusion. In stage 2, the updated and amended search was screened by one author (MW) in reverse chronological order back to 2018, and full papers checked by two other authors (NB, SI).

<u>Selection and appraisal of documents</u>

We included documents that met our definition of VWs and which focused on people with frailty, older adults and/or people with multimorbidity. The selected documents also had to provide evidence suitable for theory building. We focused on evidence from the UK, but also included a set of seminal papers from Dublin, identified early in the review, which have influenced VW development in the UK, and relate to similar population demographics.

We excluded VWs in care homes, children, people with COVID, and people with a single, specific condition (e.g., cancer, cystic fibrosis).

<u>Data extraction</u>

Data extraction for theory development was carried out in two stages. First, we extracted relevant data from an initial set of eight 'core' documents, generating 'if-then-because' statements that captured causal insights about the range of VW components; one author (NB) grouped these under component headings, representing IPTs (Appendix IV). The three authors worked closely to ensure similar understanding of concepts and coding.

Second, following the update and extended search, we extracted any new information or evidence that refuted or elaborated on the preliminary CMOCs generated from the IPTs (NB). In the light of new evidence and stakeholder discussions, one author (NB) updated/revised the CMOCs and these were checked by two authors (MW and SI), giving a final set of 12 agreed CMOCs (Appendix V).

We also collected information on all included studies, including patient details, features of VWs, and their components (MW). See Appendix VI.

<u>Stakeholder consultation</u>

We consulted with stakeholders (clinicians and public contributors) in two stages.

Stage 1: After generating the initial set of IPTs, we produced a diagram of the 'patient pathway' (Figure A1), with more detailed information in a second slide (Figure A2), all based on the intervention descriptions in the core papers. We recruited three stakeholders: one clinician (a GP who had set up a COVID VW and also was a frailty lead with Gloucestershire CCG (HL)) and two public contributors: one a carer of a relative with frailty and one who was a patient in a COVID VW.

We held two informal online meetings (Microsoft Teams), one with the clinician (facilitated by MW/SI) and the other with the two public contributors (facilitated by our PPI co-ordinator).
We presented Figure A1, Figure A2 and the list of components in the order of the patient pathway (Table A1.1 column 1), alongside frailty stability definitions from two papers [4, 5] to stimulate discussions with stakeholders. This resulted in further if-then-because statements from the meeting transcripts.

Stage 2: Following the generation of the preliminary CMOCs, the team engaged with a second group of stakeholders in six online (Microsoft Teams) meetings (two sessions with the original clinician, two sessions with a geriatrician with experience of care home VWs (TW) and one session with another geriatrician with experience of frailty VWs). We also recruited five new public contributors, one of whom had experience of VWs through her GP surgery work, and one patient who had had frailty.

We presented the new CMOC statements in a structured way to stimulate discussion, facilitated by NB/MW. These stakeholder meetings focused on aspects to prioritise, and we also compared our findings with the new NHS England guidance. [6]  Stakeholder feedback led to the extended search as above.

Synthesis and analysis

Synthesis was led by one author (NB) with contributions from two others (MW, SI).

Initially, we grouped the 'if-then-because' statements thematically under 21 intervention components. Using the intervention component as a starting point (resource), we constructed preliminary context-mechanism-outcome configurations (CMOCs), defining mechanism as 'reasoning of stakeholders in response to resources and context as the backdrop contingencies necessary for that mechanism to 'fire'.

Synthesis was iterative, and once a new CMO configuration was generated during the synthesis, we also revisited previous papers. New CMOCs were added to the list. We produced a full set of final CMO configurations, alongside the source evidence (Appendix V), and these findings were checked and agreed upon by the three authors. We did not perform rigour assessment for the data. This was because of resource constraints of the rapid review.

We grouped the final set of 12 CMOCs into three sections:

A. VW building blocks/underpinning structures, i.e., how the system context influences VW professionals.
B. VW delivering the patient pathway – key components of frailty management, i.e., how the VW context influences VW professionals and patient outcomes.
C. Patient and Caregiver Experience, i.e., how these stakeholders are included as part of the team (or not).

**Figure A1: Overview of Virtual Ward model**

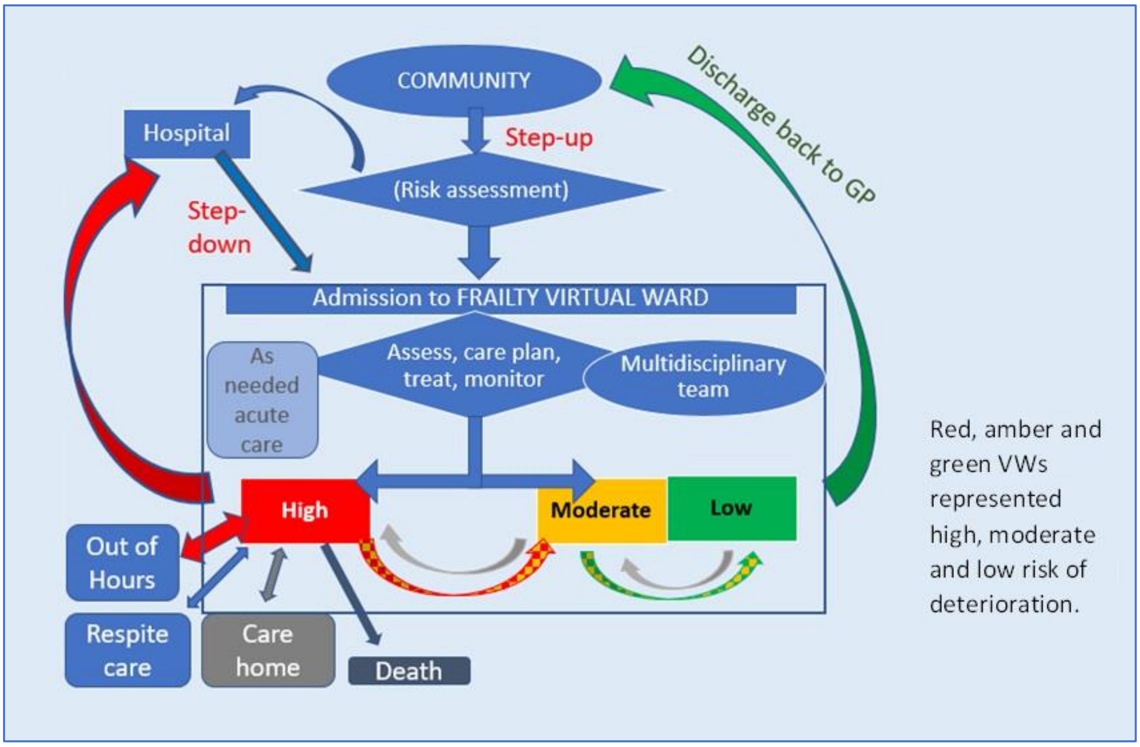

**Figure A2: Detail of Virtual Ward model**

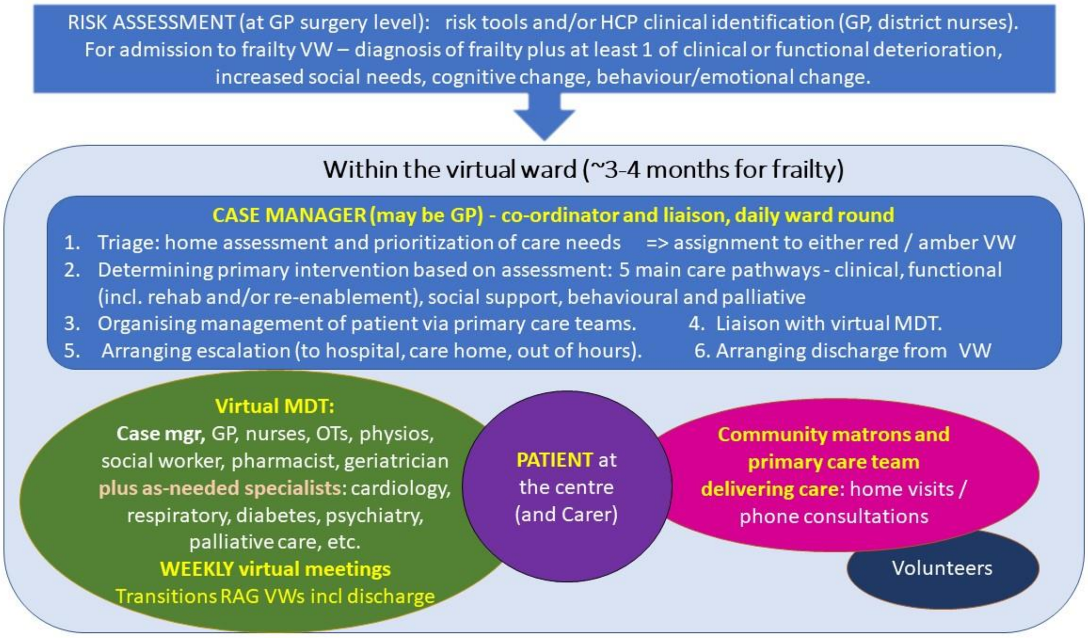

### APPENDIX II – Defining Virtual Wards

There are various definitions of VWs in the literature, with one debate revolving around whether Hospital-at-Home and VWs are the same thing. They are in essence both multidisciplinary team initiatives facilitating expert input for the patient in their own homes, reducing the need for hospitalisation for crisis management.

We had developed our definition of VWs, informed by literature identified at the scoping stage:

- Patients are offered admission to a virtual ward if a risk prediction tool identifies them as being at high risk of an unplanned hospital admission. Patients remain in the community during their time on a virtual ward, and receive multidisciplinary care intended to maintain or improve their health status and reduce their risk of unplanned hospital admission. Care is delivered in person at the patient's home, by telephone and/or at a local clinic. Virtual ward staff discuss patients on office-based 'ward rounds', participating either in person or by telephone. [7]

- All four criteria of: (1) The care provided is similar to that provided by an interdisciplinary hospital ward team, (2) Care is longitudinally coordinated by an interdisciplinary team comprising at least two health professionals (e.g. MD, Nurse); (3) Care may be delivered in the patient's home, through telephone or at a local clinic; (4) Care can include telemonitoring and case managers; however, there must be clear and evident oversight and integration of patient care by the interdisciplinary team. [8]

- Virtual wards could be said to be different to a hospital-at-home because virtual wards admit people at high risk of hospital admission (or readmission for populations post-discharge) and hospital-at-home provides co-ordinated, multidisciplinary care in the home for people who would otherwise be admitted to hospital, as a full substitute for acute hospital care. Virtual wards are a pre-emptive health intervention. [8]

- However, the model of care uses the staffing, systems and daily routines of a hospital ward to deliver preventive care to patients in their own homes in the aim of mitigating their risk of unplanned hospitalisation. [9]

- "Unlike a traditional ward being made up of beds in a physical hospital, the patients' own beds become part of a virtual ward. Patients' care remains hands-on, but it's given in the comfort of their own homes instead of a hospital. The virtual bit of a virtual ward is the way multi-disciplinary teams of health and care professionals plan each patient's care, using digital technology to help them meet". [10]

- "Why do we call it virtual? Does it involve lots of computers and technology? No this isn't the case… The virtual ward will mean that community care teams will be able to support people in their own home, providing treatment in their home environment and supporting people and caregivers so they can get better more quickly by avoiding some of the problems that can happen when people are admitted to hospital. To help community staff, such as nurses and GPs, hospital-based healthcare professionals will be available over the telephone to provide advice and support where needed.
  This is why it's called a virtual ward, as people would be supported by a wider care team including hospital staff that would be typically found on a ward. This will often mean that we can treat people at home and reduce the need for hospital care". [11]

**Our definition of VWs**

Based on this literature, we defined VWs as:

- the VW cares for patients in their own homes (in the community)   AND
- there is an MDT that makes decisions/plans patient care remotely (virtually)   AND
- the MDT provides oversight and integration of patient care

In December 2021, NHS England produced guidance on the introduction of VWs for patients with 'acute exacerbations of conditions related to frailty'. [6]  Recent work has also focused on short-term VWs or hospital-at-home models for acute care, including a rapid evidence synthesis of systematic reviews of acute VWs, hospital-at-home and remote monitoring, across all countries, [12] and the British Geriatrics Society's position paper on VWs for older people with frailty. [13]

Throughout the review process, we reflected on emerging descriptions of VWs and hospital-at-home such as:

- "Hospital at home provides co-ordinated, multidisciplinary care in the home for people who would otherwise be admitted to hospital" and "People are admitted to hospital at home assessment in the community by their primary care physician, in the emergency department or a medical admissions unit. Hospital at home may also provide hospital-level care following early discharge from hospital".  [14]
- Virtual wards deliver hospital level processes of care, enabling increased capacity for acute clinical care to be delivered in the patient's home. [15]
- Virtual wards bridge the gap between hospitals and patient's home, allowing hospital-level care including diagnostics and treatment, using many of the same staff that work in hospitals. [15]

Subsequently, we decided to include 'hospital-at-home', provided they also met our VW definition.

### APPENDIX III – Search Strategy

**Ovid Multi-file Search – Update** (27 June 2022)

**+ Forward Citation Search (Web of Science; Google Scholar)**

Search summary:

- Virtual integrated care-UK-elderly, or virtual wards-UK, n=31 [new]
- Admission-avoidance & hospital-at-home/integrated care & UK-elderly, n=112 [new]
- Forward-citation-search (nine key studies from earlier search (Nov 2021)), n=88 [new]

Total, n=231

---

**Ovid multi-file search** (27 June 2022)

**& Forward citation search (Web of Science; Google Scholar)**

Search Strategies:

Ovid APA PsycInfo <1806 to June Week 3 2022>; Embase <1974 to 2022 June 24>;
Ovid MEDLINE(R) ALL <1946 to June 24, 2022>

**Set-1: Virtual wards/virtual integrated care – elderly - UK**

1 virtual ward?.mp.

2 remove duplicates from 1

3 (virtual* adj5 integrat* adj5 (care or health* or services or model*)).mp.

4 (virtual adj5 (inter-agenc* or interagency* or interdisciplinary or inter-disciplinary or interorgani?ation* or inter-organi?ation* or inter-profession* or interprofession* or intersectoral or inter-sectoral or joint-agenc* or jointagenc* or joint organi?ation* or jointorgani?ation* or joint-profession* or jointprofession* or jointsector* or joint sector* or multi-agenc* or multiagenc* or multidisciplinary or multi-disciplinary or multi-organi?ation* or multiorgani?ation* or multi-profession* or multiprofession* or multisector* or multi-sector*) adj5 (collaborat* or commission* or coordinat* or co-ordinat* or cooperat* or co-operat* or care or delivery or healthcare or intergrat* or model* or network* or partners* or pathway? or practice* or service? or staff* or strateg* or team or teams)).mp.

5 1 or 3 or 4

6 Aged/ or "Aged, 80 and over"/ or Frail Elderly/ or Frailty/ or Health Services for the Aged/ or geriatric assessment/ or geriatric nursing/ or geriatric psychiatry/

7 aged/ or frail elderly/ or very elderly/ or geriatrics/ or gerontopsychiatry/ or geriatric care/ or elderly care/ or exp geriatric nursing/ or geriatric assessment/ or geriatric patient/ or geriatric rehabilitation/ or gerontology/

8 elder care/ or gerontology/ or geropsychology/ or gerontological counseling/

9 (aging or ageing or elder* or frail* or geriatri* or geronto* or psychoger* or geropsych* or seniors or (late* adj (life* or adulthood)) or (old* adj (adult? or age? or people? or person? or

citizen? or men or women or male? or female? or patient? or population?)) or old old or very old or senior citizen? or pensioner? or retired or retirement or care home? or nursing home?).tw.

10 ((functionally impaired or complex) adj3 (elder* or geriatri* or geronto* or psychoger* or geropsych* or seniors or (late* adj (life* or adulthood)) or (old* adj (adult? or age? or people? or person? or citizen? or men or women or male? or female? or patient? or population?)) or old old or very old or senior citizen? or pensioner? or retired or retirement or homebound* or housebound or home bound* or house bound)).tw.

11 exp dementia/ or alzheimer disease/ or (dementia or alzheimer*).mp.

12 (("65" or "69" or "70" or "75" or "79" or "80" or "85" or "90" or "95") adj years).tw.

13 (("65" or "69" or "70" or "75" or "79" or "80" or "85" or "90" or "95") adj2 old*).tw.

14 (end-of-life or hospice*).mp.

15 or/6-14

16 5 and 15

17 remove duplicates from 16

18 Homes for the Aged/ or Housing for the Elderly/ or Senior Centers/ or Homebound Persons/

19 residential home/ or respite care/ or assisted living facility/ or nursing home/ or home care/

20 residential care institutions/ or nursing homes/ or assisted living/ or group homes/ or institutionalization/

21 ((elder* or geriatri* or old* people* or psychogeri* or retirement or senior citizen? or seniors) adj3 (centre? or center? or home? or housing or facilit* or institution? or resident*)).tw.

22 ((elder* or older* or geriatri* or psychogeri* or retire* or senior or seniors) adj3 (community adj dwelling?)).tw.

23 (institutionali* adj (elder* or old* or aged or geriatri* or psychogeri* or resident* or retire* or senior or seniors)).tw.

24 "homes for old*".tw.

25 retirement commun*.tw.

26 ((care or healthcare) adj3 home?).tw.

27 ((institutionali* or resident* or respite) adj3 care).tw.

28 ((resident* or respite) adj3 (centre? or center? or facilit* or home?)).tw.

29 ((assist* or commun*) adj (dwelling? or housing or living)).tw.

30 Long-Term Care/px or Residential Facilities/ or Respite Care/ or Assisted Living Facilities/ or Group Homes/

31 ((long term or longterm) adj care adj3 (facilit* or home? or institut* or setting)).tw.

32 (communit* adj (care or healthcare*) adj (facilit* or home? or institut* or setting)).tw.

33 ((day or daily) adj3 care* adj3 (cent* or facilit* or home? or institution* or setting)).tw.

34 ((care or nursing) adj home?).tw.

35 ((discharged or posthospital* or post hospital*) and (elder* or geriatri* or geronto* or psychoger* or geropsych* or seniors or (late* adj (life* or adulthood)) or (old* adj (adult? or age? or people? or person? or citizen? or men or women or male? or female? or patient? or population?)) or old old or very old or senior citizen? or pensioner? or retired or retirement or homebound* or housebound or home bound* or house bound)).tw.

36 or/18-35

37 5 and 36

38 remove duplicates from 37

39 17 or 38

40 (UK or England or Ireland or Scotland or Wales or Britain).af.

41 (Aberdeenshire or Avon or Bristol or Bedfordshire or Buckinghamshire or Cambridgeshire or Cheshire or Cornwall or Antrim or Armagh or County Down or Cumbria or Denbighshire or Derbyshire or Devon or Dorset or Dundee or Durham or East Riding or Edinburgh or Essex or Glamorgan or Glasgow or Gloucestershire or Greater Manchester or Gwynedd or Hampshire or Hereford* or Hertfordshire or Herts or Highlands or Kent or Lancashire or Leicestershire or Lincolnshire or London or Londonderry or Merseyside or Midlands or Newport or Norfolk or Northamptonshire or Northumberland or Nottinghamshire or Oxfordshire or Pembrokeshire or (Perth and Kinross) or Shropshire or Somerset or Staffordshire or Stirlingshire or Suffolk or Sussex or Swansea or (Tyne and Wear) or Yorkshire or Warwickshire or Wiltshire or Worcester*).af.

42 (Aberdeen or Armagh or Bangor or Bath or Belfast or Birmingham or Bradford or Brighton or Bristol or Cambridge or Canterbury or Cardiff or Carlisle or Chelmsford or Chester or Chichester or Coventry or Derby or Dundee or Durham or Edinburgh or Ely or Exeter or Glasgow or Gloucester or Hereford or Inverness or Hove or Hull or Lancaster or Leeds or Leicester or Lichfield or Lincoln or Lisburn or Liverpool or London or Londonderry or Manchester or Newcastle or Newport or Newry or Norwich or Nottingham or Oxford or Perth or Peterborough or Plymouth or Portsmouth or Preston or Ripon or Salford or Salisbury or Sheffield or Southampton or St Albans or (St Asaph or Llanelwy) or St Davids or Stirling or Stoke-on-Trent or Sunderland or Swansea or Truro or Wakefield or Wells or Westminster or Winchester or Wolverhampton or Worcester or York).af.

43 (Bassetlaw or Berkshire or Black Country or Blackburn or Blackpool or Bolton or Bradford or Brent or Bury or Calderdale or Cannock Chase or Castle Point or Chorley or Coventry or Darwen or Doncaster or Dudley or Ealing or East Riding or Fareham or Farnham or Formby or Fulham or Frimley or (Fylde and Wyre) or Gateshead or Glossop or Gosport or Hackney or Halton or Hammersmith or Hardwick or Havering or Heywood or Hillingdon or Hounslow or Hove or Huddersfield or (Isle adj1 Wight) or Ipswich or Kernow or Kirklees or Knowsley or Luton or Medway or Milton Keynes or Morecambe or Newham or Preston or Oldham or Redbridge or Rochdale or Rochford or Rotherham or Rugby or Rutland or Salford or Sandwell or Seisdon or Sefton or Solihull or Southend or Southport or St Helens or Stafford or Stockport or Stoke On Trent or South Ribble or Sunderland or Surrey or Swindon or Tameside or Tees Valley or Telford or Thurrock or Tower Hamlets or Trafford or Tyneside or Wakefield or Walsall or Waltham Forest or Warrington or Waveney or Wigan or Wirral or Wrekin).af.

44 (NIHR or NHS or National Health Service).mp.

45 or/40-44

46 39 and 45

**Set-2: Virtual wards - UK**

47 2 and 45

48 46 or 47

49 remove duplicates from 48

**Hospital-at-Home**

50 ((avoid* or prevent*) adj3 (admission? or re-admission? or readmission? or admitted or re-admitted or readmitted)).mp.

51 "hospital at home".mp.

52 ((inter-agenc* or interagency* or interdisciplinary or inter-disciplinary or interorgani?ation* or inter-organi?ation* or inter-profession* or interprofession* or intersectoral or inter-sectoral or joint-agenc* or jointagenc* or joint organi?ation* or jointorgani?ation* or joint-profession* or jointprofession* or jointsector* or joint sector* or multi-agenc* or multiagenc* or multidisciplinary or multidisciplinary or multi-organi?ation* or multiorgani?ation* or multi-profession* or multiprofession* or multisector* or multi-sector*) adj5 (collaborat* or commission* or coordinat* or co-ordinat* or cooperat* or co-operat* or care or healthcare or intergrat* or network* or model* or partners* or practice* or services or staff or strateg* or team or teams)).mp.

53 50 and (51 or 52) and (15 or 36)

54 45 and 53

55 ((admission? or re-admission? or readmission? or admitted or re-admitted or readmitted) adj (avoid* or prevent*) adj (team? or model*)).mp.

56 54 or 55

57 remove duplicates from 56

58 limit 57 to yr="2017 -Current"

**Virtual Wards – Forward citation searches** 27-June-2022

| Study Reference | #1 Fwd-Cit (Web of Science) | #2 Fwd-Cit (Google-Scholar) | #1 OR #2 |
|---|---|---|---|
| Cushen B, Madden A, Long D, Whelan Y, O'Brien ME, Carroll D, O'Flynn D, Forde M, Pye V, Grogan L, Casey M, Farrell K, Costello RW, Lewis C. Integrating hospital and community care: using a community virtual ward model to deliver combined specialist and generalist care to patients with severe chronic respiratory disease in their homes. Irish Journal of Medical Science. 2022; 191(2):615-621. doi: 10.1007/s11845-021-02633-z. Epub 2021 May 6. PMID: 33956325; PMCID: PMC8100740. | 0 | 0 | 0 |
| Jefferson-Loveday, CA. A new virtual ward; assessing its impact on elderly patients in the Poole North locality in Poole, UK. Age and Ageing, 2019;48(1):i1–i15, https://doi.org/10.1093/ageing/afy211.07 | 0 | 0 | 0 |
| Jones J, Carroll A. Hospital admission avoidance through the introduction of a virtual ward. British Journal of Community Nursing. 2014;19(7):330-4. doi: 10.12968/bjcn.2014.19.7.330. PMID: 25039341. | 0 | 21 | 21 |
| Kirkcaldy A, Jack BA, Cope LC. Health care professionals' perceptions of a community based 'virtual ward' medicines management service: A qualitative study. Research in social and administrative pharmacy. 2018;14(1):69-75. doi: 10.1016/j.sapharm.2017.02.001. Epub 2017 Feb 3. PMID: 28216092. | 4 | 5 | 6 |
| Lewis C, O'Caoimh R, Patton D, O'Connor T, Moore Z, Nugent LE. Risk Prediction for Adverse Outcomes for Frail Older Persons with Complex Healthcare and Social Care Needs Admitted to a Community Virtual Ward Model. Clinical Interventions in Aging. 2020 Jun 22;15:915-926. doi: 10.2147/CIA.S236895. PMID: 32606633; PMCID: PMC7320026. | 1 | 3 | 3 |
| Lewis C, Moore Z, Doyle F, Martin A, Patton D, Nugent LE. A community virtual ward model to support older persons with complex health care and social care needs. Clinical Interventions in Aging. 2017 Jun 26;12:985-993. doi: 10.2147/CIA.S130876. PMID: 28721026; PMCID: PMC5498784. | 17 | 29 | 30 |

| | | | |
|---|---|---|---|
| Lewis G, Vaithianathan R, Wright L, Brice MR, Lovell P, Rankin S, Bardsley M. Integrating care for high-risk patients in England using the virtual ward model: lessons in the process of care integration from three case sites. International journal of integrated care.2013;13:e046. https://doi.org/10.5334/ijic.1150 | 21 | 45 | 44 |
| Lewis G, Wright L, Vaithianathan R. Multidisciplinary case management for patients at high risk of hospitalization: comparison of virtual ward models in the United kingdom, United States, and Canada. Population Health Management. 2012 Oct;15(5):315-21. doi: 10.1089/pop.2011.0086. Epub 2012 Jul 12. PMID: 22788975. | 11 | 30 | 27 |
| Stockham A. Leadership in practice: an analysis of collaborative leadership in the conception of a virtual ward. Nursing management. 2016;23(6):30-34. | 0 | 11 | 8 |
| Total | | | 139 |
| Duplicates removed within this set | | | 33 |
| Duplicates removed from other searches | | | 18 |
| Forward citations to screen | | | **88** |

### APPENDIX IV – Overview of CMOC Development

<table>
<tr><th>Original headings – stage 1 (March 2022)</th><th>Numbers of if…then…because… statements: original (and after initial stakeholders' meetings)</th><th></th><th>Initial CMOCs for discussion with 2nd stakeholders' group (June 2022)</th><th>Final CMOCs (November 2022)</th></tr>
<tr><td colspan="2">Cushen 2021; Jones 2014; Kirkcaldy 2018; Lewis G. 2013b; Lewis C. 2017; Lewis C. 2020; Lewis C 2021; Sonola 2013 (Kings Fund) [4, 5, 7, 16-20]</td><td rowspan="28"></td><td rowspan="28">• Agreeing common standards<br>• Integrated IT systems & information sharing<br>• MDT composition in the local system<br>• Learning (and sustaining) new ways of working<br>• Holistic assessment and proactive care planning<br>• Interventions at home<br>• Remote patient monitoring<br>• Caregiver's role<br>• Prioritising and selecting the right patients<br>• Patient self-management<br>• Patient discharge (or escalation)<br>• Working as a team of teams</td><td rowspan="28">• Common standards agreements<br>• Information sharing processes<br>• MDT composition and co-ordination<br>• MDT meetings<br>• Patient selection<br>• Comprehensive assessment<br>• Medication management<br>• Intensive case management<br>• Proactive care<br>• Improved communication<br>• Being at home<br>• Caregiver role</td></tr>
<tr><td colspan="2">'BEFORE'</td></tr>
<tr><td>The problem with fragmented care</td><td>9 (plus 2 clinician, plus 2 PPI)</td></tr>
<tr><td>Focus on a defined group of patients</td><td>14 (plus 1 clinician)</td></tr>
<tr><td>Instigating change</td><td>6 (plus 6 clinician)</td></tr>
<tr><td>Working as an inter-organisational, multidisciplinary team</td><td>13 (plus 12 clinician, 1 PPI)</td></tr>
<tr><td>General Practitioner Involvement</td><td>7</td></tr>
<tr><td>Using a predictive model</td><td>4</td></tr>
<tr><td>Other routes to referral</td><td>1</td></tr>
<tr><td colspan="2">'DURING'</td></tr>
<tr><td>Multidisciplinary meetings for case management</td><td>6</td></tr>
<tr><td>Inter-organisational data sharing & information management</td><td>12 (plus 1 clinician)</td></tr>
<tr><td>Shared assessments / Common standards</td><td>3</td></tr>
<tr><td>Care documentation</td><td>2 (plus 2 clinician)</td></tr>
<tr><td>Home visits</td><td>13 (plus 2 clinician, plus 2 PPI)</td></tr>
<tr><td>Patient care/communication</td><td>3 (plus 4 clinician, plus 5 PPI)</td></tr>
<tr><td>Patient safety and security (?)</td><td>0 (plus 4 clinician, plus 4 PPI)</td></tr>
<tr><td>Carer role Out-of-hours</td><td>1 (plus 1 clinician)</td></tr>
<tr><td colspan="2">'AFTER'</td></tr>
<tr><td>Patient discharge</td><td>2</td></tr>
<tr><td>VWs over time -change, sustainability</td><td>10 (plus 1 clinician, plus 1 PPI)</td></tr>
<tr><td>Avoiding hospitalisation (?)</td><td>0 (plus 2 clinicians)</td></tr>
<tr><td colspan="2">OTHERS</td></tr>
<tr><td>Negotiating contracts</td><td>4</td></tr>
<tr><td>Identifying, Labelling, Evaluating VWs</td><td>2</td></tr>
<tr><td>Post-discharge from the VW</td><td>1</td></tr>
<tr><td>TOTAL</td><td>113 literature<br>(plus 38 clinician, plus 15 PPI)</td></tr>
</table>

### APPENDIX V – Table of Context-Mechanism-Outcome Configurations

#### A) VW Building blocks

#### CMOC1: Common standards agreements

| | Context | Resource | Reasoning | Outcome |
|---|---|---|---|---|
| Common standards agreements | There is sufficient impetus and motivation in the local context towards VWs for common standards agreements to be put in place between the different providers and specialities involved in the VW, such that legal and regulatory requirements of the different authorities involved can be met. | Common standards agreements cover issues such as patient eligibility, assessment procedures, care pathways, documentation, data protection, and discharge. These standards are suitable for the working practices and cultures of different teams involved. | Having agreements in place about the purpose and processes of the VW allows for clarity and confidence in the functioning of the VW.<br><br>The people involved perceived shared goals or at least operational agreement within the VW. | Common standards agreements are implemented that formalise the collaboration and improve the communication between professionals.<br><br>This facilitates effective decision-making and case management, (leading to improved efficiency and patient outcomes/ experience.) |
| Notes | "Singing from the same hymn sheet." – agreed Standard Operating Procedures and terminology are key to clear communication for consistent care delivery.<br><br>Common standards agreements mean that VW processes are agreed and implemented consistently. Clarity around roles and expectations may help motivate teams and individuals to participate in the model.<br><br>Financial and policy motivations may vary between different professions and providers and some professionals may be accustomed to more reactive ways of working. Implementation of VWs as an integrated service is not likely to be sustainable unless contractual safeguards are in place. Aspiration to integrate care is more achievable when the people involved recognise a common goal, rather than perceiving the initiative as a top-down, 'cost-cutting' initiative. Further work could consider how perception of shared goals is achieved, but this may be helped by having a champion of the VW.<br><br>How these agreements are developed, communicated, and implemented could influence how professionals respond to them. There is a risk that standards are implemented inconsistently, for example if professionals do not feel confident in them, or not convinced of the rationale for change to the VW model. If processes are not appropriate – e.g., too unwieldy for practical use – then professionals might revert to previous ways of working.<br>By getting consensus or agreement on terms of references beforehand, a leader shares power. This leads to trust and people crossing over from their protected roles to share work towards a common goal.<br><br>Starting with a small number of patients and learning how to work in this way, finding what gaps might be missing, allowing the opportunity for people to contribute their perspectives, before expanding, may be preferable, especially for the risk averse. Rushing to set up a VW will be difficult because clinicians will be unhappy if they | | | |

| | |
|---|---|
| | perceive patients to be put at risk. Clinician recommendations were for VWs to start small and over time become more risk tolerant, and perhaps move towards more proactive care, as the model becomes established into practice. This has implications for common standards agreements – they will need to be flexible to tolerate change and will likely require frequent review. |
| Example IPT | “If working practices and culture differ between organisations, then the implementation of common standards will be hampered because it will take longer for the teams to begin to use shared assessments and care plans.” (Lewis G. 2013b)<br><br>“If the context is of insufficient staff and increased workload in general practice and the community, then staff worry that virtual wards will create additional work, leading to more stress and don’t want to change to virtual ward”. (Sonola 2013 (Kings Fund)). |
| Core evidence | (Cushen 2021; Lewis G. 2013b; Lewis C. 2017; Sonola 2013 (Kings Fund)), Clinician 1, Clinician 2 & 3. |
| Additional sources | Armstrong 2012; Baker 2016; Elston 2022; Pearson 2017; Shepperd 2022; Stockham 2016 |
| Examples | (Stockham 2016): “Cross-organisational collaboration and streamlining of services can deliver more efficient team-working, however implementing organisational change to service delivery as a contractual obligation can generate resistance to it.”<br><br>(Pearson 2017): “The backdrop to the project was one of significant pressure on resources and strained working relationships between hospital and community teams. Sources of tension included concern that service reconfiguration would intensify an already-pressured workload.”<br><br>(Elston 2022): "Beneficial organisational factors include a history of collaboration between GPs and community teams and a well-developed voluntary sector, the co-location of different professional teams enabling informal MDT working; and shared clinical leadership, supported by a GP who was also a Locality Clinical Lead (a system-wide post to support acute and primary care integration)”<br><br>(Baker 2016): “Rather than trying to start with a big bang, many of the schemes in this report have undergone a phased development. This has meant services can be built around patients and clinicians and learnings can be incorporated.” |

### CMOC2: Information sharing processes

<table>
<tr><th></th><th>Context</th><th>Resource</th><th>Reasoning</th><th>Outcome</th></tr>
<tr><td>Information sharing</td><td>There is sufficient interest in the VW model, progress towards IT integration in the local context, and trust between the different providers involved for information sharing practices to be established between the organisations involved.</td><td>New data management processes are developed and implemented that facilitate effective and real-time information sharing that provides professionals with an accurate ‘whole system’ view of the patient record.<br><br>This is likely to include alert systems with emergency and out-of-hours care.</td><td>Professionals feel confident they will have accurate information when needed.<br><br>Patients feel reassured that information is available for decision-making and appreciate not having to repeat themselves.</td><td>Patient management is improved because care decisions can be better informed and made more quickly or in a timelier fashion (ideally 24/7) so that appropriate interventions can occur promptly, and processes of care streamlined.</td></tr>
<tr><td>Notes</td><td colspan="4">Information sharing encourages trust and cooperation - we think this gives professionals more confidence in the information they have available to them, which helps to improve decision-making within a prompt timeframe, and avoids duplication of effort between settings.<br><br>Integrated IT systems or access to electronic health records provide accurate, up-to-date, and reliable information for professionals. Over time, VW partners may be granted increased access to IT infrastructure as trust in the individuals and VW model is developed. Essential that those involved have adequate IT hardware and software permissions for this information sharing to work.<br><br>Seamless information sharing improves patient perception of ‘being in safe hands’ where staff have the right information.<br><br>Requires a ‘robust IT portal’: there is a risk that IT systems do not effectively facilitate information sharing so that professionals still have incomplete or delayed access to patient information. This impedes timely decision making and interventions. professionals might then become unmotivated to participate in the VW.<br><br>Unclear whether different VW models operationalise information sharing in the same way. Bespoke/adapted processes likely to be needed in different areas.</td></tr>
<tr><td>Example IPT</td><td colspan="4">If a single IT solution is not present in a virtual ward, then case management will not be collaborative, because all stakeholders (GP, A&E, Social care) will need to record information separately and not be able to share effort, information, or decisions. If current and reliable information is available, case management will be improved because OOH decisions can be made in a timely fashion. (Jones 2014)<br><br>If there is existing infrastructure for integration in the region, then data sharing can remain contentious, because it is difficult to navigate legal requirements and guidance from different authorities. (Lewis G. 2013b)<br><br>If organisations work on different IT systems, then there is no data sharing, and often the extensive pre-setup and work-up of patients is wasted, because the emergency department starts again with patient information after deterioration has taken place. (Clinician 1)</td></tr>
</table>

| | |
|---|---|
| Core evidence | Jones 2014; Kirkcaldy 2018; Lewis G. 2013a; Lewis G. 2013b; Sonola 2013 (Kings Fund). PPI. Clinician 1 |
| Additional sources | Colligan 2015; Shepperd 2022; Stockham 2016. PPI 2 |
| Examples | (Stockham 2016): "a major weakness in the collaborative working system lay not with the working culture and individuals, but with the intersectoral technology because patient-related information had to be transferred through direct contact or secure mailing systems, subsequently affecting the working environment."<br><br>(Shepperd 2022): "integration of the electronic patient record system with GP practices was considered to have improved communication about prescriptions and reduced potential medication errors… Lack of access to information technology systems between hospital and social services was a key issue."<br><br>(Colligan 2015): "A system was put in place to 'flag' the patients on the Virtual Ward so they are easily identifiable as a Virtual Ward patient. This was vital to ensure that if a patient presented at A&E or required medical input from GP 'Out of Hours' services, the attending doctors would be aware of the patient's status and could access information regarding their ongoing care and treatment plan." |

### CMOC3: Multidisciplinary team composition and coordination

| | Context | Resource | Reasoning | Outcome |
|---|---|---|---|---|
| Multidisciplinary team composition and coordination | Frailty is a multidimensional condition shaped by unique personal circumstances of each patient. People with frailty have complex health and social care needs requiring multi-disciplinary input. The expertise required for patient management may be disparate across multiple teams. | The multidisciplinary team in the VW includes primary care, community care, and speciality frailty clinicians.<br><br>Team composition varies according to the aims of the VW and local patient needs - may include physiotherapist, pharmacist, social worker, mental health professional, voluntary sector, other clinical specialities like cardiology, respiratory, neuropsychiatry, palliative.<br><br>Team coordination is likely to be carried out by the VW coordinator(s). | Team composition and coordination encourages professionals to trust that the model can provide safe and personalised care for patients at home (and will not put patients at risk). They feel willing to participate in the model. | Team collaboration means that patient management within the VW benefits from expertise and skills from different specialisms and organisation. Team composition and coordination improves patient access to a range of interventions and support, which improves patient outcomes.<br><br>Trust within the team facilitates sharing of tasks, which can remove unnecessary steps of patient care (including travel time between patients' homes). |
| Notes | VW team composition aligned to the domains of the comprehensive geriatric assessment can facilitate appropriate multidisciplinary action. Team composition might encourage professionals to trust the VW, particularly if appropriate escalation pathways are planned for. If relevant organisations are not involved, clinicians can lack confidence in the model (e.g., option to escalate to day hospital). Avoidable delays in decision-making could be caused by gaps in team composition (e.g., access to physiotherapy).<br><br>Continuous change in the local context (e.g., due to other improvement projects, pandemics, workforce issues, etc) might contribute change weariness so that professionals find it harder to 'buy-in' to the change to the VW model or aspects of it. So while the perception of sufficient (and responsive) senior support may be important, for professionals to 'buy-in', it may also be beneficial to have key recognisable individuals that can champion the new model, especially from GPs.<br><br>Even if not explicitly involved in the VW team, co-operation with other organisations will be required for effectiveness including the voluntary community sector, ambulance service, social care, and mental health).<br><br>A VW coordinator role is usually carried out by a senior nursing/professional who plays a vital role, coordinating every part of what happens on the ward. The | | | |

| | |
|---|---|
| | coordinator needs to have a good all-round clinical background or knowledge of what is available in the community to support patients at home. The coordinator can strive towards seamless integration between organisations and avoid potential duplication of effort within services.<br><br>Sharing of tasks and training could enhance teamwork and awareness of mutual strengths and limitations, leading to a shared approach to problem solving. Enhancing integration with acute and primary care, pharmacy and the voluntary sector can lead to additional benefit for the patient and the health and social care system. Including the voluntary and community sector in the VW means that longer term or more holistic aspects can be factored in – such as social isolation. |
| Example IPT | If there is a lack of understanding of the wider perspective and aim of the virtual ward, collaboration and consequent success of virtual ward is undermined because of role protectionism in the team. (Jones 2014)<br><br>If members of the MDT want the virtual ward to succeed, then there will be a synergistic effect that benefits patients, because they are willing to work together and get involved in cases (to a greater or lesser extent). (Sonola 2013 (Kings Fund)) |
| Core evidence | Cushen 2021; Jones 2014; Lewis G. 2013b; Lewis C. 2017; Sonola 2013 (Kings Fund). Clinician 1 |
| Additional sources | Baker 2016; Elston 2022; Leeds_CCG 2019; Pearson 2017; Shepperd 2022; Swansea-Bay 2020 |
| Examples | (Leeds_CCG 2019): "The virtual ward is a consultant led service that supports people experiencing medical problems in their own home. There is rapid access to diagnostics (e.g. pathology / radiology) and treatments that can be safely delivered at home (e.g. intravenous medicines). However because it is a multiagency team including social care colleagues, people also get rapid access to increased care packages and therapy services where required."<br><br>(Elston 2022): "In this locality, the MDT comprises general practitioners (GPs) (with read and write-access to all Coastal GP records), pharmacists, and voluntary sector Well-being Coordinators in addition to the community matrons, community nurses, occupational and physiotherapists, social workers, mental health liaison staff and health and social care co-ordinators found in other localities (with external GP input requested when needed). The EIC team was co-located within the Teignmouth Health & Well-being hub (a former community hospital), where health and social care staff are jointly managed."<br><br>(Baker 2016) Midlothian: ""The service provided to patients is fully integrated across specialisms and sectors… This means care packages can be provided at home for patients who need additional care, which is one of the biggest benefits of being an integrated team providing health and social care."<br><br>(Swansea-Bay 2020): "With input from our virtual ward geriatrician, GP lead and representatives from the voluntary sector, we were able to provide wraparound support for the patient and family, easing their concerns"<br><br>(Swansea-Bay 2020): "The co-ordination of the team and the support involved is essential and part of my role is to feed back and be a point of contact for the family should they have concerns. I believe having that link and support at the end of the phone has made this experience a positive one and has improved the care for this patient." Quote Team co-ordinator<br><br>(Shepperd 2022): "Failing to integrate with longer-term services, such as district nursing, could be a problem. Professionals highlighted the need to manage the rising demand for domiciliary or social care in the context of cuts in state funding, as this is a key constraint to implementing health policy that is aimed at reducing hospital admissions." |

| | |
|---|---|
| | (Shepperd 2022): “Staff considered that sharing traditional disciplinary roles could enhance teamwork by increasing awareness of mutual strengths and limitations… … Extended scope training of nurses, physiotherapists, occupational therapists and pharmacists, and at one site also of paramedics, was undertaken to share approaches to problem-solving and ensure that a common language was used that would enable interprofessional communication in the team.“<br><br>(Pearson 2017): “Inter-professional working was facilitated through a focus on joint decision-making, co-location of rehabilitation staff, and an emphasis on implementing changes through consensus… Supportive relationships across teams, united by the strategic vision provided by Geriatrician involvement, provided the framework on which referral and information-sharing processes could be built. These supportive relationships also enabled practitioners to feel more secure in moving services towards a pro-active, patient-centred, therapy-led approach.“ |

### CMOC4: Multidisciplinary team meetings

| | **Context** | **Resource** | **Reasoning** | **Outcome** |
|---|---|---|---|---|
| Multidisciplinary team meetings (*capacity / shared learning)* | The aims of the VW model and its implementation facilitates (and ideally motivates) different teams and disciplines to work together, and the team members have sufficient capacity to attend and participate in regular multidisciplinary team meetings. | The multidisciplinary team meets regularly, either in-person or via technology, to discuss patients.<br><br>The meetings provide a forum for communication between specialist clinicians and the care teams providing hands-on care.<br><br>VW coordination helps the meetings to run smoothly. | Professionals perceive MDTs to be effective and worthwhile.<br><br>Better communication and shared learning encourage the development of collaborative relationships.<br><br>Effective teamwork enables professionals to feel more secure in the VW model. | Communication within the MDT facilitates holistic patient care and prevents avoidable delays in decision-making.<br><br>Meetings improve patient management by enhancing the effectiveness and efficiency of decision-making.<br><br>Further, effective MDT meetings could allow for collaborative leadership, role sharing, staff empowerment, and upskilling/role development. |
| Notes | MDT meetings enable the VW to function as a forum for the integration and prioritisation of patient care. In these meetings, the MDT discusses patients with a frequency depending on their needs.<br><br>The multidisciplinary teamwork in a VW allows experts and generalists to work together to provide holistic patient care -meetings of the multidisciplinary team provide a hub for decision-making in the VW. If professionals engage with MDT meetings and build trusting relationships there will be better communication and more collaborative working.<br><br>The mix of specialist skills is very important, but frequent meetings held remotely from the patient might be unattractive to staff who value face-to-face human connection.<br><br>Sufficient clinical capacity is required to make it happen (and keep it going) – competing workloads could make it hard for professionals to work together, especially if professionals are not given protected time for the VW work.<br><br>The technology gives a route for meetings to occur much more easily, which is especially valuable when teams are already stretched or there is distance to travel.<br><br>Meetings could allow for shared learning amongst the team, but there is less evidence on how to run these meetings to maximise this shared learning. It is unclear how conflict is managed/decision-making resolved, whether authority defaults to 'senior' staff, and how operating remotely impacts on relationship development and depth of learning as a team. There is a risk that the management style or processes of implementation impedes team communication.<br><br>Meetings could be less effective without consistency of core VW staff or if too few patients are admitted to the ward. Perceived disparity in attendance at MDTs could influence motivation of others to engage, eventually leading to a regression to old ways of working. | | | |

| | |
|---|---|
| | VWs might work best when there is diminished formal role distinction and more sharing of tasks, so that activities that are less professionally segregated, and cohesive teamwork is encouraged. This is important for efficiency when working across a geographical area. Where there is an awareness of mutual strengths and benefits and an educational focus on role development, knowledge can be exchanged beyond traditional disciplinary boundaries which could strengthen a shared learning experience and improve perceptions of different professional groups involved. |
| Example IPT | If regular multidisciplinary team meetings occur and are attended by professionals from community healthcare, primary care, and social care, then care integration and communication between providers can be improved and shared values and trust can be fostered, because there is a forum in which patient care can be discussed. (Lewis G. 2013b)<br><br>If there is a perceived disparity in GP attendance at virtual ward meetings, then other members will feel disinclined to attend and the MDT will be less effective. (Sonola 2013 (Kings Fund)) |
| Core evidence | Jones 2014; Lewis G. 2013b; Sonola 2013 (Kings Fund) |
| Additional sources | Baker 2016; BNSSG_CCG 2020; Elston 2022; Pearson 2017; Rankin 2010; Shepperd 2022; Stockham 2016 |
| Examples | (Elston 2022): Co-location of the different teams involved in the virtual ward, or with in the key external organisations, could confer a level of connectedness that supports joint working.<br><br>(Shepperd 2022): If parts of the system are not visible to each other, this could further service fragmentation because it impacts on collective understandings of how to meet frailty needs at home.<br><br>(Pearson 2017): “The backdrop to the Improvement project was one of significant pressure on resources and strained working relationships between hospital and community teams. …There was suspicion about the way that any service reconfiguration would impact on workload, work scheduling, and expectations about responsibilities"<br><br>(Rankin 2010): “The virtual ward provides a positive forum and opportunity to improve and build on positive professional relationships and assist in our joint working. I believe it is mutually supportive in approach and it is informative in helping each of us to understand how the different agencies represented at the meetings are organised, their different pressures and from which perspective we approach our work. This in turn assists us in working more productively together for the benefit of the service users we serve.”<br><br>(Stockham 2016). “All participants were encouraged to communicate their skills and thereby establish their individual boundaries. This process not only produced clarity but also created an air of empowerment. Collaborative leadership and management structure improved the working culture. In breaking down professional barriers, a more cohesive workforce evolved.”<br><br>BNSSG (BNSSG_CCG 2020) Sirona health director quote: "The Virtual Ward Round consists of highly skilled multi-disciplinary professionals and is supporting our clinicians to help more service users receive the right care and support; it is also proving to be a useful sharing of information and learning environment for those involved.” |

### B) VWs delivering the patient pathway

#### CMOC5: Patient selection

| | Context | Resource | Reasoning | Outcome |
|---|---|---|---|---|
| Patient selection | Frailty is a multidimensional condition with interacting facets, in which functional or clinical decline is often precipitated by cognitive or physical deterioration.<br><br>GPs (as part of the GP contract) are required to use frailty risk tools and therefore have appropriate information to identify people in the community who are in crisis or nearing a 'tipping point' into crisis (at high risk of frailty-related deterioration or hospitalisation).<br><br>There are limited NHS resources for VWs, so there is a need for patient selection and prioritisation. | Patient selection processes provide GPs (possibly alongside or in addition to others such as community matron and secondary care consultants) with a referral route that can improve the management of complex health and social care needs related to frailty (ideally before a deterioration becomes a crisis).<br><br>Patient selection processes therefore provide the professionals in the VWs with a prioritised group of patients on whom to focus their efforts (and a clear rationale for doing so). | Professionals perceive that they can have an impact in keeping these patients safe and preferably at home, and so make the effort to work together. | The patients who are selected benefit from a period of proactive multidisciplinary input that provides timely/early interventions tailored to their needs. The input might stabilise their frailty and prevent a crisis, thus reducing the risk of unplanned hospitalisation and/or length of stay if admitted. |
| Notes | Effective patient selection/referral routes could be critical to VW function for there to be sufficient patient referrals and for the patients who are referred to be appropriate. Perceptions that the VW is prioritising and selecting the 'right' patients (e.g., taking an acceptable stance on risk of harm) may be important for the buy-in from professionals, and to patients and their families or caregivers. Effectiveness of patient selection will be shaped by common standards agreements made during the implementation of the VW (e.g. the extent to which referral criteria 'work' in practice).<br><br>Referral should take account of whether the intervention can be effective in helping that patient (not just divert the 'difficult-to-manage'). Perception of the potential impact of the VW might be influenced by the referring professional's knowledge and experience of VWs and its team composition.<br><br>Referral might be informed by predictive risk modelling and be based on other criteria including frailty severity scores and clinical judgment. The effectiveness of a predictive model would rely on the completeness of its data input, the ease with which clinicians could interpret its outputs, and for those clinicians to have capacity and willingness to | | | |

| | |
|---|---|
| | do so. Risk prediction tools based on solely on hospitalisation may be less suitable in frailty and should also include frailty severity and impactability. |
| Example IPT | “If virtual wards target patients at high risk of future hospitalisation that are likely to respond to the proposed intervention, then they can reduce unplanned hospitalisations because they provide a focus for the integration of care.” (Lewis 2013) |
| Core evidence | Lewis 2013, Lewis 2017, Kings Fund 2013, Clinician 1 |
| Additional sources | Baker 2016; Colligan 2015; Leeds_CCG 2019; Sheffield 2018; Shepperd 2021; Swansea-Bay 2020), Clinician 3 |
| | (Colligan 2015): reports that using algorithmic patient selection was time-consuming and inefficient – instead, patients were identified by GPs, Allied health professionals, district nurses, and social care staff.<br><br>(Sheffield 2018): “For GP practices, a virtual ward provides a consistent, proactive approach to caring for people with the most complex medical and social needs in the community, rather than a reactive one that could end up in multiple hospital admissions.”<br><br>(Baker 2016) Midlothian “In many cases the GP simply does not have the time to sort out the complex issues, and they [GPs] have found the [virtual ward] service very helpful in providing more intensive support for their patients."<br><br>(Baker 2016) South Sefton: “Most staff have access to a common IT platform. The virtual ward screens patients for medication issues, falls, dementia and nutritional status. The community matron meets regularly with the GPs in her allocated practices. Together they identify older people with frailty who are at high risk of a crisis, in order to enrol them in a programme that can last up to three months.”<br><br>(Shepperd 2021): “We recruited older people with frailty who required an urgent hospital admission because of an acute change in their health, such as a sudden functional deterioration, delirium, or a fall, against a background of complex comorbidity… Most participants were referred from an acute assessment unit or an older persons’ frailty unit, with only a minority referred directly from home by their GP”. |

### CMOC6: Comprehensive assessment and evaluation

| | Context | Resource | Reasoning | Outcome |
|---|---|---|---|---|
| Comprehensive Geriatric Assessment (or alternative holistic frailty assessment) and generating a shared Care and Support Plan | Frailty needs are multidimensional - recommended treatment for frailty is the Comprehensive Geriatric Assessment (CGA).<br>Shared assessment processes and care documentation are agreed and implemented within the VW.<br>Team composition and the functioning of the MDT facilitates access to interventions, specialists, and services so as to be responsive to the multiple domains of the CGA. | Patients are assessed by a VW coordinator using a suite of screening tools (e.g. as part of the CGA).<br>Face-to-face contact and the holistic assessment identify psychological, environmental, and social needs associated with potential frailty events.<br>The coordinator then works with the MDT to prepare (and enact) a tailored Care and Support Plan. | Professionals in the MDT feel confident in the information available from the assessment.<br>The VW co-ordinator feels confident in the support they receive from the MDT.<br>Patients feel peace of mind from a comprehensive assessment and continued communication with the VW co-ordinator. | Comprehensive assessment and subsequent care and treatment identifies and resolves immediate clinical concerns and potentially vulnerable areas.<br>Appropriate professionals are mobilised according to individual patient needs, such that patients receive timely access to specialists and interventions which improve their outcomes.<br>Reduced duplication of effort (compared with 'silo-ed' care) in assessment and treatment may prevent avoidable delays in access to interventions; and improve patient and staff satisfaction. |
| Notes | In most cases it appears that the VW co-ordinator carries out initial assessment such as the CGA or an assessment based on this and drafts a tailored case management plan with the patient/caregiver, then leads on enacting that plan with the rest of the multidisciplinary team.<br>A comprehensive assessment process contributes to the more cohesive approach to case management, which can then inform proactive care planning. The assessment and care and support plan includes a plan for monitoring progress. Patients can be triaged within the VW into red, amber, and green VWs, which determines the frequency of monitoring or of review by the MDT.<br>We think a consistent point of contact is reassuring for patient and caregiver, and that improved communication with them means their needs and preferences are more likely to be met. We are not sure what happens if the VW coordinator is indisposed. Patients appreciate not having to repeat themselves with successive assessment processes. However the use of new assessment processes may be hindered if they are perceived as unwieldy or too divergent from existing working practices. | | | |

| | |
|---|---|
| Example IPT | If a suite of screening tools (e.g. Frailty index, pressure ulcer risk, activities of daily living, cognitive levels, etc), is used to assess dependency and care, then patients will be less likely to experience deterioration or hospital admission, because their care needs will be identified quickly, they can be risk-stratified, and appropriate targeted interventions given.<br><br>(Lewis C. 2017): If virtual wards can do initial problem solving, then patients are less likely to have hospital admission, because of some gain in health related outcomes relatively quickly, and becoming equipped to self manage. (Lewis C. 2017)<br><br>If, following assessment, patients in the Virtual Ward are subdivided according to need and care plan into "daily" beds (red), "weekly" beds (amber) and "monthly" beds (green), then optimum care across the virtual ward can be achieved, because the frequency with which different patients are reviewed on a ward round can be determined such that the MDT can spend most time on the patients at greatest risk. (Lewis G. 2013a) |
| Core evidence | Cushen 2021; Lewis G. 2013a; Lewis C. 2017 |
| Additional sources | Colligan 2015; Shepperd 2022 |
| | (Colligan 2015): "Similar to a hospital ward manager, the Case Co-ordinator was responsible for co-ordinating the case and clearly communicating with all involved, to ensure a seamless integrated service and avoid potential duplication of services… The virtual ward does not just look at the chronic condition: the co-ordinator spotted a suspicious lesion on one of my patients and correctly identified it as malignant – the patient received timely intervention."<br><br>(Shepperd 2022): Staff considered that undertaking assessments in a patient's home could enhance their awareness of safety factors when compared with the limitations of assessments in hospital… The relevance of CGA was disputed among staff at another site, with some identifying its importance and others expressing the view that full CGA was not feasible as part of acute assessments." |

### CMOC7: Medication management

<table>
<tr><th></th><th>Context</th><th>Resource</th><th>Reasoning</th><th>Outcome</th></tr>
<tr><td>Medication management by a dedicated team</td><td>Polypharmacy is common in people with frailty, because of its multi-dimensional nature.<br><br>Specialist input for medication management can enable complete and accurate medication reviews.</td><td>VW includes (or has access to) a dedicated medication management team or expertise.<br><br>Conducting a personalised medication review in the home setting facilitates accurate medication reconciliation that is not always possible in an outpatient clinic.<br><br>Unnecessary polypharmacy can be identified and resolved with the wider expertise of the MDT</td><td>The VW team are better informed, and if the reasons for medicine review are explained to the patients/ caregivers, they would feel supported and ideally more educated about their medicines.</td><td>Reduced polypharmacy could lessen the burden of treatment and improve the effectiveness of medication.<br><br>Reduced side effects and risk of adverse events could improve patient outcomes and reduce the cost of pharmaceuticals in some cases.<br><br>Treatment adherence may be improved where patients are better informed.</td></tr>
<tr><td>Notes</td><td colspan="4">Specialist medication review within the VW reduces the need for medication review by GPs or other professionals – but only to the extent that those professionals trust the information provided by the medication management team.<br><br>There is a risk that the medication management team do not have enough time or skills for accurate medication reconciliation and regime modification in the particular patient context (e.g., memory loss).<br><br>‘Joined-up’ communication with the patient and caregivers about medication can resolve confusion and increase their understanding. They may then feel better able to manage because some of the symptoms of polypharmacy are removed, and VW staff can address any fears. In contrast, unexplained removal of medications may be a source of anxiety.</td></tr>
<tr><td>Example IPT</td><td colspan="4">If a dedicated med management team is included within a virtual ward they may reduce polypharmacy and the need for reviews by the GPs and specialists because the team have the time and skill to do a review of medication for the patients at their homes and can modify regimens. They lead to better care because they discuss with patients their medication which increases their understanding around their meds thus increasing adherence. However they may not lead to better care when patients have complex medical conditions and memory and mental health issues because the team do not have enough time and skills needed to deliver the service to these patients. (Kirkcaldy 2018)</td></tr>
<tr><td>Core evidence</td><td colspan="4">Cushen 2021; Kirkcaldy 2018</td></tr>
<tr><td>Additional sources</td><td colspan="4">Colligan 2015; Shepperd 2022. Clinician 2, PPI 2</td></tr>
<tr><td>Examples</td><td colspan="4">(Colligan 2015): “The Virtual Ward co-ordinator fully assessed Mr W optimising medication and providing education to develop him as an expert patient. Mr W and his wife both report a significant improvement in quality of life and are both more confident in dealing with the exacerbations which are part of his chronic lung condition. GP feedback: ‘Excellent service, it’s great that the co-ordinator can fully assess the patient and prescribe appropriate medication without the GP having to visit. I have been kept fully informed of the care prescribed’."<br><br>(Shepperd 2022): “Communication about changes to medication following discharge from either hospital or HAH could be a problem”</td></tr>
</table>

### CMOC8: Intensive case management

<table>
<tr><th></th><th>Context</th><th>Resource</th><th>Reasoning</th><th>Outcome</th></tr>
<tr><td>Intensive case management and monitoring of people with frailty</td><td>Frailty and multi-morbidities can mean complex health and social care needs as a result of rapidly fluctuating or deteriorating health.</td><td>Regular MDT meetings allow for intensive and integrated case management.<br><br>Regular in-person visits and (optionally) remote monitoring increases the frequency and quality of contact between the patient and the VW.</td><td>The staff providing hands-on care, and the MDT making remote decisions, feel informed about when to modify treatment.<br><br>Patients/caregivers feel 'visible' to the health and care system in a way that feels safe and supported.<br><br>It is possible that patients/ caregivers have reduced anxiety and improved understanding of symptoms (e.g., better able to recognise adverse symptoms).</td><td>Treatment can respond rapidly to changing patient need and interventions can be delivered within the patient's home.<br><br>Patients can be safely managed because clinical deterioration is rapidly detected and acted upon in a timely manner, which prevents further decline/ escalation because early interventions and short-term responses can be arranged at home.<br><br>Monitoring and review also allow the MDT to determine when a patient is stable and ready for discharge</td></tr>
<tr><td>Notes</td><td colspan="4">Monitoring provides the team with reliable information so that they feel well-informed on when to step-up and step-down care, and ideally there is enough face to face contact that patient/caregivers feel safely supported.<br><br>VWs improve access to appropriate and timely support by identifying patients to prioritise, determining their needs, and helping them 'jump the queue' to get those needs met.<br><br>Following assessment and throughout the duration of the stay in the VW, the patients can be triaged by severity and acuity into red/amber/green VWs, which determines the frequency of monitoring and review by the MDT and also the type of treatment (e.g., patients in a red VW may initially have acute care (such as subcutaneous fluids and intravenous antibiotics) then moving to proactive care. It is possible that a 'traffic light' system helps patients, caregivers, and clinicians be aware of and encouraged by progress.<br><br>If there is deterioration, patients can contact the VW co-ordinator during working hours and systems are in place to allow rapid access to out-of-hours / emergency services. This gives 24/7 contact with the 'VW'.<br><br>For remote monitoring, patients (and caregivers) need to be able and willing to agree to some level of technology use. However, their context confers different capabilities for technology – including Wi-Fi availability.</td></tr>
</table>

| | |
|---|---|
| | There is a risk that remote monitoring and intensive case management increases, rather than decreases, patient anxiety and so does not encourage their self-management or increases the caregiver burden.<br><br>There may be a range of criteria and different experiences for discharge from the VW into the community, including: formal discharge policies, MDT decision making criteria, continuity of care plans, post-discharge review or poor communication and a lack of information. In some VWs, patients and caregivers may be uncertain when discharge will occur and professionals may not agree over the relevance of the CGA in short-term wards. |
| Example IPTs | If the virtual ward framework allows for the monitoring of patient symptoms and prioritization of care needs, then patients will feel more confident about remaining at home, because additional support services can be mobilized in a timely manner. Red-flag clinical presentations will be identified, and early interventions given, as well as specialist follow-up. (Lewis C. 2017)<br><br>If, following assessment, patients in the Virtual Ward are subdivided according to need and care plan into "daily" beds (red), "weekly" beds (amber) and "monthly" beds (green), then optimum care across the virtual ward can be achieved, because the frequency with which different patients are reviewed on a ward round can be determined such that the MDT can spend most time on the patients at greatest risk. (Lewis G. 2013a)<br><br>If there is a cohesive approach to case management and decision making, then patients can be appropriately discharged to usual care provided by the primary care team, because proactive care planning and the ability to determine when a patient is stable are possible within the virtual ward (Lewis C. 2017)<br><br>If there is no virtual ward, then the patient is more likely to go into hospital because there is no identification of problems and no timeliness of decision making (Caregiver 1) |
| Core evidence | Jones 2014; Lewis G. 2013a; Lewis C. 2017; Sonola 2013 (Kings Fund), Clinician 1, PPI 1 |
| Additional sources | Baker 2016; Rankin 2010; Shepperd 2022, PPI 2 |
| Examples | (Shepperd 2022): “Consultants were readily accessible during normal working hours and in a way that they would not necessarily expect in a hospital setting. They [People delivering care at home] attributed their confidence in facing unpredictable conditions while undertaking home visits, sometimes in remote locations, to having reliable support, often in the form of rapid telephone access to senior practitioners at the team base."<br><br>(Rankin 2010), Patient’s wife: “Very happy with the service provided. It has really made a difference to us, not just in better health but also in sorting out outpatient appointments, booking transport and being able to take blood samples at home.”<br><br>(Shepperd 2021): "Employing remote monitoring alongside multi-disciplinary care might also have a role but would have to be balanced against the care needs of this population."<br><br>(Shepperd 2021): “Many who received HAH described not knowing how long to expect the service to be available or had not anticipated imminent discharge: That just came out the blue”<br><br>(Shepperd 2021): “the relevance of CGA was disputed among staff at another site, with some identifying its importance and others expressing the view that full CGA was not feasible as part of HAH acute assessments. Team members at this site considered the purpose of CGA to be enabling the patient’s medical condition to be stabilised at home, if possible, and then referring the patient to community rehabilitation or other services, if required.” |

### CMOC9: Proactive care

| | Context | Resource | Reasoning | Outcome |
|---|---|---|---|---|
| Proactive care and enablement | Frailty and co-morbidities can mean fluctuating or sudden/rapid deteriorations in health.<br><br>Patient (and caregivers) have a positive attitude towards self-management and anticipatory care, such that they are able to participate in proactive care planning and implementation.<br><br>Care pathways / anticipatory interventions have sufficient capacity for patients to be seen in a timely manner. | Management of frailty includes preventative or anticipatory measures and in some cases the patient being taught strategies to self-manage.<br><br>Patients and caregivers receive holistic input and potentially a new or updated home care package (i.e., with support for hydration, nutrition, and personal care).<br><br>Other primary or community interventions might include mental health, advanced care planning, falls prevention, physiotherapy, social support | Professionals and patients / caregivers perceive that potential issues can be addressed or avoided instead of or prior to escalation.<br><br>Patients/caregivers feel safe and supported, ideally more able to cope, and more confident in managing at home.<br><br>Patients and caregivers who understand proactive care have improved confidence and feel empowered through the process. | Patients receive care and other interventions in their homes, tailored to their needs.<br><br>Improved quality of life and patient safety/ satisfaction.<br><br>Advance planning means decisions can be made in advance of a crisis that help the patient to avoid hospital.<br><br>Lower risk (but not zero risk) of adverse events leading to hospitalisation.<br><br>Discharge out of the VW to the care of the GP. |
| Notes | Proactive care is prevention rather than reacting when something goes wrong. It is intended to get treatment in place before the situation reaches a crisis point and to help stop things becoming worse, for the patient and their family as well<br><br>VW holistic assessment includes both personal care needs and proactive care planning so that support can be reviewed / delivered within the VW. Ideally patients receive rapidly responsive and proactive interventions in their home, tailored to their needs, to prevent adverse events and stabilise frailty. That could include arranging or updating a home care package and other proactive interventions including psychological and physiotherapy. The existing relational resources of family, the neighbourhood and community professionals in the VW can be built on to act as a bridge to continuity of health care.<br><br>A proactive and holistic approach to care includes an element of anticipatory planning, but it is unclear extent of advanced care planning in different VWs. Care coordination / advance planning means that decisions can be made in advance that help the patient to avoid hospital. Patients or caregivers that do not have sufficient capacity to participate in this might feel less confident and might not feel listened to during communication with the VW. These patients do not engage with what is offered by the VW and their caregivers could then experience additional stress.<br><br>So ideally patients (and caregivers) feel better equipped to self-manage. However, patients without the awareness, knowledge, skills, and confidence to manage their own healthcare could find it challenging to become 'active' in their care. There is a risk | | | |

| | |
|---|---|
| | that health anxiety could be exacerbated in a way that limits empowerment towards self-care, and possibly then fosters dependency within the VW. (There is little evidence on patient/caregiver experience within VW to unpack this more.) |
| Example IPT | If patients have positive thinking about achieving self-care goals, then they gain maximum benefits from the virtual ward, because they take responsibility for their care and engage with the anticipatory care offered in the virtual ward. (Jones 2014)<br><br>If virtual ward members (e.g. GP, paramedics, frailty nurses, pharmacists) collaborate [both] together and with the patient on the virtual ward, then preventative measures can be put in place in advance of a crisis (e.g. RESPECT forms sorted). (Clinician 2022) |
| Core evidence | Jones 2014; Lewis C. 2017; Sonola 2013 (Kings Fund), Clinician 1 |
| Additional sources | Armstrong 2012; Baker 2016; Shepperd 2022; Swansea-Bay 2020, PPI 2 |
| Examples | (Baker 2016) South Sefton; GP: "We weren't really making much progress. But the virtual ward holistic care has really given [the patient] the skills to manage her conditions better. She comes into my room now with a smile on her face."<br><br>(Shepperd 2022): "Discussions with patients differed from those in hospital settings, as non-medical practitioners would undertake complex discussions, for example about end-of-life care. This difference was portrayed positively, as non-medical staff would have spent time in the home and could discuss issues in a timely and responsive way as they arose."<br><br>(Armstrong 2012): "Our current systems and services do not offer the right quality. We have services which have grown historically and in an unplanned way; become poorly aligned with the needs of local patients; high levels of variation from area to area; silos, leaving gaps in care pathways; duplicate process, such as assessments; too many 'hand-overs' of care, which generate confusion amongst patients and clinicians…. Our current systems and services are also too reactive and hospital-centric. This is not affordable, doesn't offer good quality care for patients and is not sustainable as our population changes." |

### C) Patient and Caregiver Experience

#### CMOC10: Improved communication

| | Context | Resource | Reasoning | Outcome |
|---|---|---|---|---|
| Improved communication between the VW and patient/ caregiver, and with out-of-hours/emergency services. | VW processes ensure that communication and information sharing with the patient/caregiver is well established and can continue out of usual working hours.<br>This means that the VW patient can contact a professional 24/7; and that alert systems are in place to notify the VW if patients have contact with emergency care/out of hours. | Ready access to the VW co-ordinator and having 'out-of-hours' contact mechanisms improve communication between the patient/caregiver and the VW team, and between HCPs, which reduces the time taken to seek and receive assistance. | Personal contact with the care co-ordinator, who the patient knows, is reassuring.<br>Patients, caregivers, and professionals trust that the necessary information will be available for decision-making in the event of clinical fluctuation or crisis.<br>With more awareness of the support in place, patients/caregivers may feel reassured and less vulnerable. | Patients and professionals can receive timely and accurate information.<br>Improved communication facilitates holistic and responsive care.<br>Improved communication with the patient and caregiver means their needs and preferences are more likely to be met, and anxiety may be reduced. |
| Notes | Improved communication can help to avoid hospitalisation because there is a route to seek help when health is deteriorating.<br>Treatment adherence and general wellbeing can be improved.<br>Being in the home environment, but still feeling visible to healthcare in an emergency, requires improved communication routes. Few VWs have 24 hour cover.<br>Risk that patients/caregivers feel more vulnerable when returning to normal contact mechanisms (e.g., to GP). Ideally there should be clear communication around discharge expectations.<br>Patients might have remote and automated monitoring as well as face-to-face, but there is not much evidence on this within frailty populations. Patients may be happy with technology for monitoring or may find it to be a barrier. | | | |
| Example IPT | If the virtual ward can act as a link between various care providers, then communication can be better, and confusion minimised (Kirkcaldy 2018)<br>If there is personal contact, then some patients will be reassured, because there is someone talking them through it (PPI 1)<br>If the virtual ward has no face to face contact, then patients will lack confidence in the model, especially if they live on their own (Caregiver 1) | | | |
| Core evidence | Kirkcaldy 2018; Lewis G. 2013b; Lewis C. 2017, Clinician 1, PPI 1, Caregiver 1 | | | |
| Additional sources | NHS_Wales_Award 2015; Rankin 2010; Ryland 2015; Sheffield 2018; Shepperd 2022; Swansea-Bay 2020, PPI 2 | | | |
| Examples | (Sheffield 2018): "What are the benefits of virtual ward to a person? Not having to repeat your story. Having the same team of professionals involved in your care who | | | |

| | |
|---|---|
| | know what 'well' looks like to you personally. Choosing to be part of the virtual ward means that 'true' person-centred care can be delivered. So for example when Derek has a flare-up of his respiratory problems and has trouble talking, another healthcare professional can see from his 'OK to stay' care plan that this is 'common' for Derek and given a 'bit of time' and one of his inhalers, he can safely stay and be treated at home where he prefers, rather than go into hospital unnecessarily."<br><br>(Ryland 2015), Patient Feedback: "The virtual ward team were only a phone call away, that was good knowing that someone was there when you need help." "The care I received from the team was first class. When you live alone, nights can be frightening, but knowing I could get in touch 24 hours made me feel safe. Thank you all."<br><br>(Swansea-Bay 2020): "The co-ordination of the team and the support involved is essential and part of my role [case manager] is to feedback and be a point of contact for the family should they have concerns. I believe having that link and support at the end of the phone has made this experience a positive one and has improved the care for this patient."<br><br>(Rankin 2010)- Patient: 'Normally I would have called 999 for an ambulance but the doctor came out, prescribed a 3 day course of steroids and I didn't have to go to hospital'.<br><br>(NHS_Wales_Award 2015), Patient: "The nurse came in and she said, 'Nothing to worry about – I'll give you a telephone number - never mind what time of day or night it is for me'. So that gave me peace of mind".<br><br>(Shepperd 2022): "Communication about changes to medication following discharge from either hospital or HAH could be a problem"<br><br>(Shepperd 2022): "At no site did HAH staff routinely provide copies of discharge summaries to patients, although staff said that their final discussions with patients should involve talking through the discharge plan and any medication changes. However, some patients felt that there was a lack of information, and some sought advice from their GP: there was no guidance e.g. on cutting down painkillers" "Some patients/caregivers interviewed reported a lack of clarity about the timing of discharge from HAH, and a lack of involvement in planning for discharge" |

### CMOC11: At home instead of hospital

| | Context | Resource | Reasoning | Outcome |
|---|---|---|---|---|
| Being [safe] at home | People with unstable frailty may need to attend hospital during a health or care crisis, but extended or repeated stays in hospital may impact negatively on the health/wellbeing of a person with frailty (and potentially their families/informal caregivers).<br><br>In general, patients may prefer to be at home where possible, rather than in hospital. | The VW facilitates integrated case management and appropriate interventions to support people with frailty in their own home – through rapid access to MDT input, diagnostics, and treatments (and in some cases, social care and other therapeutic/ rehabilitative services). | Patients and caregivers feel comfortable and secure at home and happier, in a familiar home setting.<br><br>They feel supported to stay at home safely and reassured by having the communication routes to the VW.<br><br>(However, some patients / caregivers may not feel confident or safe in their own homes.) | Appropriate and timely interventions are delivered to the patient at home, improving or stabilising their condition, supporting activities of daily living, and reducing risk of hospitalisation.<br><br>Staying at home allows continuation of established routines (inc. mobility) and existing varied forms of support (e.g., neighbours). |
| Notes | Ideally, people with frailty are treated proactively in the home to prevent frailty-related crises and consequent hospital admission. People have rapid and better care and treatment as needed in VW, because the VW has a multiagency team that includes diagnostics, therapy, and social care all-in-one. The VW provides more optimal support to the patient because multiple visits from varied sources can be made for different needs at their homes; in some VWs this is day or night. The patient's home environment can be unpredictable, so staff making home visits should have appropriate support to deal with non-routine events or challenging situations.<br><br>Continued self-management at home could be made more likely, to the extent that the home environment enables established routines in a familiar setting to continue, potentially with some modifications. Ideally after a VW stay, patients are thus enabled to manage better at home; in contrast, after a hospital stay, caregivers might not have confidence that they can manage the patient at home because this represents a large change from hospital. Being at home also means that family/friends do not have to travel to/visit the patient in hospital.<br><br>Most VWs do not provide a 24/7 service and rely on caregiver support and contact with out-of-hours/emergency services. This can be a source of concern. For some patients/families the hospital is a safe environment that confers peace of mind - the home environment feels less 'safe' during a crisis. There may also be reasons why a patient's home environment is not suitable for the interventions to be delivered (e.g., smokers at home – oxygen tanks). Patients without any informal caregiver support or who have a lot of health anxiety may need special consideration.<br><br>Safety, especially in short term wards is often maintained by families, who may not be able to contain risks at home. Depending on the patient's condition, home may feel unsuitable and unsafe, particularly if the patient has acute confusion or falls.<br><br>Note that in some emergencies, hospital would be the appropriate answer and so VWs may not avoid hospitalisation entirely but could reduce length of stay if | | | |

| | |
|---|---|
| | admitted. PPI members expressed that they didn't want to be 'an elderly bed-blocker', and so they would be grateful for the VW if it stopped them getting in that position. |
| Example IPT | If there is a combined approach to care, with home visits and follow-up telephone consultations, then patients' quality of life will be improved because patients and caregivers feel supported at home (Lewis C. 2017) |
| Core evidence | Cushen 2021; Jones 2014; Lewis C. 2017; Sonola 2013 (Kings Fund), PPI 1 |
| Additional sources | Colligan 2015; Leeds_CCG 2019; NHS_Wales_Award 2015; Shepperd 2022; Swansea-Bay 2020, PPI 2 |
| | (Leeds_CCG 2019): "The virtual ward is a consultant led service that supports people experiencing [acute] medical problems in their own home. There is rapid access to diagnostics (e.g. pathology / radiology) and treatments that can be safely delivered at home (e.g. intravenous medicines). However because it is a multiagency team including social care colleagues, people also get rapid access to increased care packages and therapy services where required. People can be supported at home with multiple visits through the day and care overnight if needed. Their care plan will be reviewed daily by the virtual ward MDT meeting."<br>(Colligan 2015): "There are occasions where to maintain a patient at home, a Social Care Domiciliary Care package requires to be commenced or increased."<br>(Shepperd 2022): "It is possible that the patient's independence is maintained by the recovery in the familiar home setting.. Many patients considered their home a place of familiarity and security but, for some, it had also become a place of vulnerability, which could have implications for families…<br>(Shepperd 2022): Safety was often maintained by families, with family caregivers temporarily moving into the patient's home or family caregivers arranging for the patient to move into the caregiver's home.<br>(Shepperd 2022): "Patients and caregivers recognised acute health care in the home necessitated their involvement in monitoring safety: 'It's like sleeping with one eye open, it's almost like sleeping with one ear open.' "<br>"For some, continuity through community services became particularly important in regaining confidence"<br>(Shepperd 2022): "However, it can be precarious for families to contain risks at home, e.g. a mother did not recognise her son at night and tried to get out of the window… Those living separately from the patient were particularly concerned about the lack of 24-hour care…. sometimes people just need to get themselves better in hospital, to have all the treatment and have the 24-hour care that they have there, which they wouldn't have at home".<br>(Shepperd 2022): "Staff described situations when patients would be excluded from HAH, sometimes but not always because of the absence of a caregiver at home or being alone at night and there were concerns about safety…. The assessment of safety to be particularly difficult when patients were experiencing acute confusion or had been falling." |

### CMOC12: Caregiver experience

| | Context | Resource | Reasoning | Outcome |
|---|---|---|---|---|
| Caregiver's role (partner / family / friends etc.) | Family or informal caregivers are impacted by a person with frailty's fluctuating health.<br><br>Caregivers provide additional practical and emotional support to patients, and navigate health care and social systems to support usual routines | VWs facilitate integrated care, timely interventions, and proactive decision-making.<br><br>(Where appropriate), caregivers may be included by the VW coordinator in communication about the patient and shared decision – making, and this gives the VW more insight on the patient's situation. | The caregiver feels reassured and supported by the VW and may perceive the responsibility for care is shared or removed.<br><br>Through being involved in decision-making during assessment and treatment, caregivers may gain knowledge that helps them feel more confident in continuing to manage in the future. | Caregiver burden and stress is reduced, which prevents burnout or other risks of harm.<br><br>Medical interventions or professional care at home could also allow the caregiver to maintain their social support networks during an acute health episode |
| Notes | Some, but not all, VWs include a partnership approach with patients and caregivers. There is less information on the caregiver's role and involvement in the VW and how it might reduce or increase their stress. It is possible that frequent contact with VW reduces anxiety and monitoring improves understanding of symptoms (e.g., better able to recognise adverse symptoms). Caregivers described awareness of subtle changes when maintaining support at home, such as recognising delirium as a symptom of an undiagnosed UTI). Adverse events might also be prevented by avoiding caregiver burnout.<br><br>VWs might only be safe for some patients if a caregiver is available – otherwise, the VW won't be suitable for patients for whom safety in their own home is at risk. Informal caregivers are expected to take some responsibility for patient safety, which could add to caregiver burden. This is especially the case if the VW is not as responsive during out-of-hours.<br><br>Some caregivers would respond differently to monitoring and frequent visits. Ideally effective care coordination reduces their stress, but there is a risk that might not feel listened to by the MDT. For example, consent to visit might be sought from the patient but not obtained from the people they live with.<br><br>Assessments should not be confined to the patient's health condition, without regard to caregivers' health needs. Enablers of caregivers in VW are listed as: their knowledge and confidence; them having a flexible, layered social network able to mobilise to meet changing needs and gaps; their involvement in VW assessment and discharge planning that enables shared decision-making; and continuity in communication with (and by) professionals, including community relationships.<br><br>Patients and caregivers often make joint decisions about how best to manage an acute health event, and these are shaped by their relationship with professionals, their social networks, and a desire to avoid a stay in hospital. For example, many were familiar with the triage and advice line NHS 111 and had used it to access immediate guidance before making direct contact with the health services. | | | |

| | |
|---|---|
| | There may be limited opportunities in short-term VWs for caregivers discussing how to manage beyond the acute episode.<br><br>Experiences of discharge can vary. One caregiver reported the best experience because of aftercare and continuity with the GP. Others said the timing of discharge may be unclear and there may be a lack of guidance; patients/caregivers may not be involved in planning. |
| Example IPT | "If the carer knows that there is a multidisciplinary team in place who are reviewing the patient, then they may have more confidence in continuing to manage the patient at home. There could be a reduction in ED presentations/admissions because caregiver burnout has been prevented." (Brainstorming meeting) |
| Core evidence | Lewis C. 2017; Sonola 2013 (Kings Fund), PPI 1 |
| Additional sources | Shepperd 2022; Vaartio-Rajalin 2019; NHS Wales Award 2015 |
| Examples | Shepperd 2022: "Many caregivers reported that the rationale for some decisions had been unclear and attributed this to the perceived lack of opportunity to convey their opinions about cognitive, communicative, and physical functioning [of the patient]"<br><br>(Shepperd 2022): "Patients and caregivers commented that HAH care was often confined to the patient's presenting health condition and that assessments did not include broader challenges, such as caregivers' health needs…. Our findings show that caregivers' capacity to provide additional practical and emotional support, and a suitable home environment, is crucial."<br><br>(Vaartio-Rajalin 2019): "Both the pre-admission phase and actual care period seem to include a focus on the patient only. During the referral process and the initial visit to the patient's home, verbal informed consent was sought from the patient but not the patient's near ones [i.e. caregivers and family]... Patients are involved in in the evaluation of care, but near-ones are not involved."<br><br>(Shepperd 2022): "Caregivers described limited opportunities for discussing with HAH or hospital staff how to continue to manage beyond the acute episode, or 'what I can do to change, if anything, the conditions of what Mum's living with' "<br><br>(Shepperd 2022): One caregiver said, "This [HAH] has been the best hospital experience from other times because there seems to be aftercare. . . normally you'd have to phone your doctor and go through whole loop again."<br><br>(NHS_Wales_Award 2015), Caregiver (husband): "When she came home from hospital well more or less, I had to take over as the heart specialist and all the responsibility. But then Eira came as a district nurse, she said 'Don't worry, Mr D. We'll do everything. And we will look after you'. And this has been the best thing that I could have had because it lifted everything off my shoulders.<br>"If we were left alone there's a possibility the <u>two</u> of us could have gone back into hospital, not the one, the two of us, because I'm nearly 76 and it could have put a lot of strain on me and with Sybil struggling as well."<br><br>(Shepperd 2022): Caregivers reflected on the unstable trajectory of the older person's health needs, and many considered that proactive reviews would be useful after discharge. Many, from both health care settings, commented on the lack of a written record that could support them to assess change"<br><br>(Shepperd 2022): Family caregivers often played a crucial role in monitoring their relative during an episode of hospital-at-home care and integrating transitional care arrangements into longer-term strategies."<br><br>(Shepperd 2022): The importance of health-care professionals' understanding of caregivers' challenges is widely established, yet their contribution to managing older people's acute health care at home is not always recognised. |

### APPENDIX VI – Included Studies

| Study detail | Virtual Ward type | Population | Intervention | Monitoring, self-management | Outcomes/aims | CMOC addressed |
|---|---|---|---|---|---|---|
| **ACADEMIC PUBLISHED LITERATURE** | | | | | | |
| Cushen 2021 [16]<br><br>CORE PAPER<br><br>Sept – Nov 2020 (i.e. during COVID)<br><br>Dublin, Eire<br><br>Proof-of-concept initiative case study | Model 2 (closest)<br><br>Urgency: both acute and non-acute (mild-mod exacerbations of respiratory conditions and stable patients with poorly controlled disease).<br><br>Duration in VW: 5-24 days<br><br>Open 24/7? – no (7-day service from 8 am to 8 pm. Out-of-hours support, and local emergency services after 8pm) | 20 patients with respiratory conditions, very mild frailty. 55% referred for disease optimisation, 40% had a current exacerbation of airways disease. Co-morbidity – median 4.5 (5.5) additional diagnoses; average 6.8 (5.3) medications per person<br><br>Selection: Initial referral criteria: patients experiencing a mild-moderate exacerbation of their underlying respiratory disease (confirmed COPD and/or asthma). Later extended to stable patients with poorly controlled disease i.e. = 2 community treated or = 1 hospital treated exacerbation in the previous 12 months.<br><br>Stratification: NR | Personalised management plan, medication reconciliation, social determinants of care, anxiety management techniques, inhaler technique, education; remote monitoring (spirometry, heart rate, oxygen saturation) | Monitoring: remote monitoring technology which facilitated monitoring of daily oxygen saturations, heart rate, and spirometry measurements, and face-to-face. All patients agreed to home visits and use of technology to remotely monitor their clinical status<br><br>Self-management: NR | Outcomes/aims: to improve clinical outcomes for patients with chronic respiratory disease. Avoid hospital admission | CMOC1 (standards), CMOC3 (MDT), CMOC6 (assessment), CMOC7 (medication), CMOC11 (at home not hospital) |

| Study detail | Virtual Ward type | Population | Intervention | Monitoring, self-management | Outcomes/aims | CMOC addressed |
|---|---|---|---|---|---|---|
| Elston 2022 [21]<br><br>2015-2018 quantitative study; 6 week qualitative study in 2017<br><br>Torbay and S Devon | Model 1b (closest)<br><br>Urgency: unclear, in-crisis and proactive care; average duration of episode 7-9 days, but may be up to 6 weeks<br>Duration in VW: unclear. Average duration of episode 7-9 days, but may be up to 6 weeks and mentions 12 weeks coaching and emotional support<br>Open 24/7? - not mentioned | Deteriorating frail older people.<br><br>Selection: Clinical referral from GPs (37.5%), from community services (25.0%) and from the acute hospital (16.7%). Two-thirds of referrals were for poor mobility or falls (40.3% and 29.2% respectively). The rest covered a range of: medical (dementia, UTIs, other) (15.3%); environmental issues, and transitions between services. GP referrals included a slightly greater proportion of medical and mental health issues (22.2%)<br>Stratification: implied (MDT functioning for in-crisis) but unclear | Reactive and proactive.<br><br>Enhanced integrated care e.g. MDT including GPs, pharmacists, and voluntary sector well-being coordinators in addition to the community matrons, community nurses, occupational and physiotherapists, social workers, mental health liaison staff and health and social care co-ordinators | Monitoring: NR<br>Self-management: Yes (goal setting tools) | Outcomes/aims: Increase service efficiency, reduce acute attendances, and provide benefits across the care system, whilst delivering a person-centred service.<br>One aim is to reduce informal caregiving and short and long-term residential care placements | CMOC1 (standards), CMOC3 (MDT), CMOC4 (MDT meetings) |

| Study detail | Virtual Ward type | Population | Intervention | Monitoring, self-management | Outcomes/aims | CMOC addressed |
|---|---|---|---|---|---|---|
| Jones 2014 [17]<br><br>CORE PAPER<br><br>2011 launch<br><br>Wyre Valley, Worcestershire, England<br><br>Historical observational study | Model 1a<br><br>Urgency: non-acute<br><br>Duration in VW: 3 months.<br><br>Open 24/7? – MDT access 7 days/week (not 24/7) | From 12 GP surgeries serving a population of 112,000 ≥1 chronic conditions<br><br>Selection: risk tools for UHA; GP referral<br><br>Stratification: informal, according to need | Proactive: assessment and anticipatory care plan and proactive treatment | Monitoring: proactive<br><br>Self-management: yes, important | Outcomes/aims: prevent admission | CMOC2 (IT), CMOC3 (MDT), CMOC4 (MDT meetings), CMOC6 (assessment), CMOC8 (intensive management, esp discharge), CMOC9 (proactive and empowering), CMOC11 (safe at home) |
| Kirkcaldy 2018 [19]<br><br>CORE PAPER<br><br>May- June 2015 (pre-COVID)<br><br>South Sefton, NW England<br>Qualitative study | Model 1a<br><br>Urgency: non-acute (implied)<br><br>Duration in VW: NR (implied longer-term).<br><br>Open 24/7? – unclear | Polypharmacy and older people with longer-term conditions implied<br><br>Selection: NR<br><br>Stratification: NR<br>Medicine management team members, 27 MDT members | Unclear, but includes medication review | Monitoring: NR<br><br>Self-management: NR | Outcomes/aims: NR | CMOC2 (IT), CMOC7 (medication), CMOC10 (Communication) |
| Lewis C. 2017, 2020, 2021<br>[4, 5, 18]<br><br>CORE PAPER<br><br>Nov 2014 – Nov 2015 (pre-COVID)<br><br>Dublin, Eire | Model 1b<br><br>Urgency: non-acute<br><br>Duration in VW: 3-7 months.<br><br>Open 24/7? – Unclear, but refers to | People with severe/moderate frailty; Rockwood CFS: 6.7 (SD 0.86)<br><br>N=54 (2017 paper); N=88 (2020 and 2021 papers)<br><br>Selection: Frailty + evidence of deterioration – Rockwood, | VW model set up to work within existing resources assessment; proactive care planning plus cognition and social support; monitoring | Monitoring: proactive, combination of face-to-face and telephone calls monitoring<br>Self-management: not a focus | Outcomes/aims: prevent unplanned hospital admission and ED presentations; achieve frailty stability; discharge from VW | CMOC1 (standards), CMOC3 (MDT composition), CMOC5 (patient selection), CMOC6 (assessment), CMOC8 (intensive management), CMOC9 proactive care, CMOC10 |

| Study detail | Virtual Ward type | Population | Intervention | Monitoring, self-management | Outcomes/aims | CMOC addressed |
|---|---|---|---|---|---|---|
| Historical observational study; [4, 18] Risk prediction study [5] | clinical assessment out-of-hours | then determine level of frailty and acuity of event. Convenience sample, referral from consultant geriatrician, following the MDT assessment, from the day hospital, outpatient gerontology clinics, or prior to hospital discharge.<br><br>Stratification: formally into red/amber/green VW | | | | (improved communication), CMOC11 (safe at home), CMOC12 (caregivers) |
| Lewis G. 2013 [7, 9, 22]<br><br>CORE PAPER<br><br>May 2006 (Croydon), Oct 2008 (Devon), March 2009 (Wandsworth) (pre-COVID) Croydon, Devon, Wandsworth, England<br><br>3 case studies, [7] non-randomised matched comparative study with non-VW controls [9] | Model 1a<br><br>Urgency: non-acute<br><br>Duration in VW: several months.<br><br>Open 24/7? – no (describes links with out-of-hours) | Selection: risk tools for UHA<br><br>Stratification: red/amber/green, according to need and care plan into “daily” beds (red), “weekly” beds (amber) and “monthly” beds (green). | Proactive; preventative care.<br><br>Individual Clinical Management Plans with goals etc | Monitoring: Face-to-face (no tele-health devices); no detail, but proactive and frequency depends on case-by-case<br><br>Self-management: mention of ‘activated’ patients | Outcomes/aims: unplanned hospital admissions; bed days; A&E admission | CMOC1 (standards), CMOC2 (IT), CMOC3 (MDT composition), CMOC4 (MDT meetings), CMOC5 (patient selection), CMOC6 (assessment), CMOC8 (intensive management), CMOC9 proactive care, CMOC10 (improved communication) |

| Study detail | Virtual Ward type | Population | Intervention | Monitoring, self-management | Outcomes/aims | CMOC addressed |
|---|---|---|---|---|---|---|
| Pearson 2017 [23]<br><br>April 2011 to March 2014<br><br>Exeter | Model 2 (closest)<br><br>Urgency: appears to be non-acute (or step-down)<br><br>Duration in VW: 6-15 days<br><br>Open 24/7? - NR (but mentions 7 days) | Older frail people<br><br>Selection: Acute Community Team referrals for all patients aged 80 years or over who were acute medical admissions to the city hospital. Single point of access for referrals, joint geriatrician and community rehabilitation practitioners review for both admission avoidance and early supported discharge<br><br>Stratification: NR | Unclear details Plan-Do-Study-Act cycles to re-reconfigure and implement a Hospital at Home service.<br>Joint geriatrician and community rehabilitation practitioners review, MDT (unclear how operated), extended weekday and weekend working hours | Monitoring: NR<br><br>Self-management: NR | Outcomes/aims: To create a sustainable comprehensive community based Hospital at Home service for older people to enable appropriate admission avoidance and early supported discharge.<br>Outcome measures: Discharge destination; Length of stay; Acute Community Team referrals. | CMOC1 (standards), CMOC3 (MDT), CMOC4 (MDT meetings) |
| RAND 2012 [24]<br><br>Autumn 2009 – Spring 2011 (pre-COVID)<br><br>Virtual wards in Sunderland, Cockermouth, Nene (Northampton-shire), Norfolk, Nottinghamshire<br><br>Series of case studies | Model 1a (closest)<br><br>Urgency: non-acute<br><br>Duration in VW: NR (implied longer-term).<br><br>Open 24/7? – NR | Older people with frequent hospital admissions (Sunderland); long-term conditions (Cockermouth); chronic conditions, older patients, and those at risk of hospital admission (Nene); vulnerable and older people (Norfolk); complex chronic care needs (Nottinghamshire)<br><br>Selection: combination of clinical decision and predictive tool<br><br>Stratification: NR | Proactive (personalised care plans, medicines management, intensive, proactive care) | Monitoring: NR<br><br>Self-management: Yes (in Cockermouth only) | Outcomes/aims: prevent hospital admission | CMOC1 (standards) and CMOC2 (IT) |

| Study detail | Virtual Ward type | Population | Intervention | Monitoring, self-management | Outcomes/aims | CMOC addressed |
|---|---|---|---|---|---|---|
| Shepperd 2021, 2022 [25, 26]<br><br>Feb 2015 to June 2018 (i.e. pre-COVID)<br><br>Multicentre study in UK:<br>Wales - Newport and Torfaen.<br>Scotland - Argyll; Livingston; Kirkcaldy.<br>England - Bradford; Exeter; London.<br>Northern Ireland - Craigavon; Belfast<br><br>Randomised controlled trial | Model 2<br><br>Urgency: acute<br><br>Duration in VW: average 7.2 (SD 5.6) days of HAH.<br><br>Open 24/7? – No (care 7 days per week 9am to early evening, admissions restricted to Mon-Fri; 24h emergency care with emergency services) | 1055 older people with frailty who required an urgent hospital admission because of an acute change in their health, such as a sudden functional deterioration, delirium, or a fall, against a background of complex comorbidity<br><br>Selection: clinical referral (patients referred by their GP to a single point of access, or who were transferred from the emergency department to an acute assessment unit and were assessed as suitable for HAH (VW)).<br><br>Stratification: NR | Reactive and proactive: geriatrician-led multidisciplinary admission avoidance HAH with CGA (including MDT and VW rounds): clinical assessments; reactive treatment – acute medical care (e.g. iv drug administration, oxygen therapy, frailty management (CGA) | Monitoring: face to face; no mention of remote monitoring<br><br>Self-management: "Team supports older person and caregiver with self-management and prevention" (no more details) | Outcomes/aims: outcomes: primary – living at home at 6 months; mortality at 6, 12 months; new long-term residential care at 6, 12 months; delirium; cognitive impairment; ADL; readmission or transfer to hospital; QoL (EQ5D); length of stay (VW or hospital); patient satisfaction; resource use; adverse effects | CMOC1 (standards), CMOC2 (IT), CMOC3 (MDT), CMOC4 (MDT meetings), CMOC5 (patient selection), CMOC6 (assessment), CMOC7 (medication), CMOC8 (case management), CMOC9 (proactive care), CMOC10 (communication), CMOC11 (at home), CMOC12 (caregiver) |
| Sonola 2013 (Kings Fund) [20]<br><br>CORE PAPER<br><br>2008 – 2012 (i.e. follow-on from Lewis 2013) (pre-COVID)<br><br>S Devon and Torbay, England | Model 1a<br><br>Urgency: non-acute<br><br>Duration in VW: several months.<br><br>Open 24/7? – no, out-of-hours clinicians working between 6pm and 8am on weekdays, | Patients with complex needs, including older people (over 65s) with several long-term conditions (majority); also a growing number of patients are in their 40s and 50s with mental health illness alongside drug/alcohol misuse.<br><br>Selection: Devon Predictive Model: high risk of | Proactive (and reactive): Assessment, triage, care plan, proactive management implementation by care co-ordinator, VW team, intermediate care | Monitoring: proactive – management depends on patient need. Face-to-face (during crisis) and telephone calls<br><br>Self-management: patient | Outcomes/aims: to identify people at risk of unnecessary hospital admissions and employ a multidisciplinary approach to address their individual needs across health and social care to prevent crises from occurring. The MDT seeks to | CMOC1 (standards), CMOC2 (IT), CMOC3 (MDT), CMOC4 (MDT meetings), CMOC8 (case management), CMOC9 (proactive care), CMOC11 (at home), CMOC12 (caregiver) |

| Study detail | Virtual Ward type | Population | Intervention | Monitoring, self-management | Outcomes/aims | CMOC addressed |
|---|---|---|---|---|---|---|
| Case study | and at weekends and public holidays. | hospitalisation in the next 12 months<br><br>Stratification: red/amber/green | team and care agencies | empowerment not evident | reduce duplication, improve continuity and the quality of care across providers and ensure that resources in the community are used efficiently. | |
| Stockham 2016 [27]<br><br>CORE PAPER<br><br>2013-16 (pre-COVID)<br><br>SE Powys, Wales<br><br>Case study and narrative discussion | Model type 1 t<br><br>Urgency: non-acute<br><br>Duration in VW: NR<br><br>Open 24/7? – NR | Chronic conditions + frailty<br><br>Selection: moderate to high-risk individuals, with chronic conditions, increased frailty and decreasing function<br><br>Stratification: NR | unclear, mentions patients remaining at home “during acute medical scenarios and chronic or frailty crises” and “unscheduled care”, but also aims for a “greater focus on anticipatory care” | Monitoring: NR<br><br>Self-management: NR | Outcomes/aims: main aim to prevent avoidable hospital admissions; also patients’ holistic management, health and wellbeing, active rehabilitation.<br>Overall to provide a greater focus on anticipatory care, thereby reducing the number of un-planned admissions | CMOC1 (standards), CMOC2 (IT), CMOC4 (MDT meetings) |
| Vaartio-Rajalin 2020 [28]<br><br>May 2019<br><br>London (Lambeth and Southwark)<br><br>Audit and interview (qualitative) | Model 2<br><br>Urgency: acute health care (instead of inpatient)<br><br>Duration in VW: NR<br><br>Open 24/7? - no, 8am-11pm 7 days/week. | Heart failure, COPD, pneumonia, cellulitis, urinary tract infections, resolving delirium, dehydration, hyperemesis, medication titration and blood monitoring. Not paediatric, psychiatric and gynaecology patients.<br><br>Selection: Referrals from hospitals and community- | Appears to be reactive only. Referral, assessment in patient's home, care plan (reviewed during MDT meetings led by consultant geriatricians at least three times per week). Daily | Monitoring: No, virtual and/or digital devices not used in patient care<br><br>Self-management: patients are involved in the evaluation of | Outcomes/aims: to facilitate early discharge from local hospitals and prevent avoidable hospital admissions by means of person-centred care based on a clinical review (status and patient's situation) | CMOC12 (caregiver) |

| Study detail | Virtual Ward type | Population | Intervention | Monitoring, self-management | Outcomes/aims | CMOC addressed |
|---|---|---|---|---|---|---|
| | | based health practitioners, including London ambulance service, district nurses and GPs. Two clinical nurse specialists are employed as a hospital-based in-reach team, and they work closely with ward and A&E teams to identify patients suitable for early discharge<br><br>Stratification: NR, but 2 VWs, each with 3 separate MDT meetings | MDT meetings and co-ordinator. | care, 'near ones' not involved | | |
| **GREY LITERATURE** | | | | | | |
| Armstrong 2012 [29]<br><br>expected future implementation 2012-2014<br><br>West Sussex, England | Urgency: may be both - proactive care in community (not urgent) and short term rapid response<br><br>Duration in VW: planned to be both short term and implied long-term<br><br>Open 24/7? - Implies not 24h | 'Frail/elderly people'<br><br>Selection: Appears to be two entry levels proposed: proactive community care and admission avoidance approaches. In the former, risk stratification (using hard AND soft intelligence). In the latter, single point of access and rapid assessment (e.g. urgent GP home visits requests seen within an hour)<br><br>Stratification: NR (except for above separation) | Reactive and proactive. Proposed proactive community care: including risk stratification; active case management; integrated MDT which will plan, coordinate and deliver care, support the whole pathway; integrated long | Monitoring: Proactive community care: proposed Assistive Technology (Telecare/telehealth), alongside other care<br><br>Self-management: Vision to enable maximum number of people | Outcomes/aims: Aim to reduce non-elective admissions, reduce length of stay, realise a significant reduction in people in long-term residential placements. Whole system approach including: proactively identifying and supporting frail/elderly people and their carers who are at the greatest | CMOC1 (standards), CMOC9 (proactive) |

| Study detail | Virtual Ward type | Population | Intervention | Monitoring, self-management | Outcomes/aims | CMOC addressed |
|---|---|---|---|---|---|---|
| | | | term condition and dementia care; Assistive Technology (Telecare/telehealth); integrate and enable proactive End Of Life care.<br><br>Proposed admission avoidance care including: CGA; rapid assessment; rapid response MDT providing acute care. | to self manage at all levels of need | risk to prevent deterioration, and avoiding all inappropriate admissions to hospital by providing CGA in the community alongside safe, robust community care. | |
| Baker 2016<br><br>(RCGP case study South Sefton, England) [30]<br><br>From 2013 | Model 1b and Model 2 (two types of VW)<br><br>Urgency: (1) VW - non-urgent and (2) urgent care team Duration in VW: up to 3 months (VW) and short-term for urgent care team (target response time is within two hours, but most cases are usually reviewed within an hour). | Majority of older adults with frailty in the area are female, have multiple long-term conditions, live alone, and require support with personal care.<br><br>Selection: Two types of programme: (1) 'virtual ward' recruits older people with frailty at high risk of a crisis (2) urgent care team - GP referral of people with frailty who would be admitted to hospital ('sub-acute') - may | Proactive (VW) and reactive (urgent care team).<br>VW (longer term) screens patients for medication issues, falls, dementia and nutritional status, proactive, responsive and holistic model to meet patients' | Monitoring: For urgent care team, staff are equipped with tele-video technology for remote assessment and support, and also face-to-face. For VW team, unclear, but assumed face-to-face | Outcomes/aims: to provide care closer to home for mild to moderate illness in older people with frailty, and care for end of life patients in their usual place of residence. A secondary goal was a reduction in unplanned hospital attendances and admissions. To facilitate coordinated | CMOC1 (standards), CMOC3 (MDT), CMOC4 (MDT meetings), CMOC5 (Patient selection), CMOC8 (case management), CMOC9 (proactive) |

| Study detail | Virtual Ward type | Population | Intervention | Monitoring, self-management | Outcomes/aims | CMOC addressed |
|---|---|---|---|---|---|---|
| | Open 24/7? - NR (but urgent care team refers to out-of-hours). | not have virtual MDT<br><br>Stratification: NR | needs; MDT and co-ordinator. The urgent care team operates out of a community walk-in centre. Tele-video technology for remote assessment and support | Self-management: NR | care across organisational boundaries, to fill the gap in community urgent care and instil a culture of continuous improvement. | |
| Baker 2016<br><br>(RCGP case study, Midlothian, Scotland) [30]<br><br>From summer 2014 | Urgency: acute care<br><br>Duration in VW: around 8 days<br><br>Open 24/7? - Not sure: but has provided care seven days a week since October 2015 | Requiring acute care. majority of patients are elderly and frail. Many have significant comorbidities such as heart or renal failure, and dementia and delirium are common. In 2015, acute infection (20%), cardiac problems (12%) and respiratory illnesses including COPD (12%). Many patients have multiple issues, such as falls, delirium and musculoskeletal problems.<br><br>Selection: GPs make the majority of referrals to the team (84% in 2015). Some patients are also referred from the local emergency department and others from the 'Front Door Geriatrician' | Reactive. Episode of specialist care delivered at home as an alternative to being treated in an acute hospital. Service provided to patients is fully integrated across specialisms and sectors. Medical input by the consultant geriatrician is provided through six sessions per week. | Monitoring: NR<br><br>Self-management: NR | Outcomes/aims: to provide more intensive support for their patients over a short time period | CMOC3 (MDT), CMOC5 (patient selection), CMOC8 (case management) |

| Study detail | Virtual Ward type | Population | Intervention | Monitoring, self-management | Outcomes/aims | CMOC addressed |
|---|---|---|---|---|---|---|
| | | in the Medical Admissions Unit of the local teaching hospital. In 2015, 15% referred by acute hospitals to support early discharge.<br><br>Stratification: NR | | | | |
| BNSSG CCG 2020 [31]<br><br>started July 2020 (during COVID)<br><br>North Somerset, England<br><br>News item | Model 2<br><br>Urgency: step-up mainly and step down (not in-crisis)<br><br>Duration in VW: not stated – within day assessment (likely short-term).<br>Open 24/7? – NR | Frailty<br><br>Selection: Seems to be clinical referral (by GPs); people with frailty<br><br>Stratification: NR<br><br>Numbers not reported, apart from 13 patients in the first week | Appears to be reactive then proactive: bespoke care plan, rapid assessment, care in community (including social care) | Monitoring: NR<br><br>Self-management: NR | Outcomes/aims: improving patient care and reducing the need for people to be admitted to hospital | CMOC4 (MDT meetings) |
| Colligan 2015 [32]<br><br>Pilot 1 Jan – 31 Mar 2009, then Jan 2010 rollout.<br><br>Cost analysis Mar 2010 to Mar 2013 (i.e. pre-COVID)<br><br>N. Ireland (North Down, Ards and Lisburn)<br><br>Case study | Model 2<br><br>Urgency: acute exacerbation, but apparently also non-acute care<br><br>Duration in VW: seems to be short term (9 and 5 days depending on year).<br><br>Open 24/7? – no (out-of-hours mentioned) | 421 patients over 3 years<br><br>Diagnosis of one or more chronic conditions (and 1 admission in past year); not necessarily frailty<br><br>Selection: high risk of hospital admission; later clinical assessment<br>Stratification: Yes depending on clinical condition | Reactive and proactive: assessment, personalised care plan, intensive support (nursing and social care), discharged when condition stabilised | Monitoring: telehealth access is available<br><br>Self-management: Yes (Expert patient programmes were encouraged) | Outcomes/aims: hospital admission, attendance at A&E | CMOC2 (IT), CMOC5 (patient selection), CMOC6 (assessment), CMOC7 (medicines management), CMOC11 (at home) |

| Study detail | Virtual Ward type | Population | Intervention | Monitoring, self-management | Outcomes/aims | CMOC addressed |
|---|---|---|---|---|---|---|
| Leeds 2020; Leeds CCG 2019 [11, 33]<br><br>Operational from Sept 2020 (during COVID)<br><br>Leeds, England<br><br>Case study | Model 2<br><br>Urgency: acute and some non-acute (step-up/step-down)<br><br>Duration in VW: up to 7 days.<br>Open 24/7? – Yes (but only taking referrals between 9am and 4pm) | Moderate or severe frailty ("co-ordinated rapid care to people in their own homes who are experiencing, or have experienced, falls, immobility, incontinence, side effects of medication or increased confusion e.g. infection, breathlessness, and are living with moderate or severe frailty")<br><br>Selection: Frailty (Rockwood or eFI)<br><br>Stratification: NR | Reactive and proactive (reactive – "treatments that can be safely delivered at home (e.g. intravenous medicines)"), but also aims to be proactive and Care Plan review and access to increased care package | Monitoring: NR<br><br>Self-management: Yes (mentioned) | Outcomes/aims: prevent hospital admission, improved patient outcomes, prevent adverse effects in hospital | CMOC3 (MDT), CMOC5 (patient selection), CMOC11 (at home) |
| NHS Wales Award 2015 [34]<br><br>Powys, Wales<br><br>2015 video case study | Model type unclear<br><br>Urgency: step-down non-acute<br><br>Duration in VW: NR.<br>Open 24/7? – Y (patient said she could phone anytime, day or night) | Older person with multimorbidities<br><br>Selection: NR (discharge from hospital)<br><br>Stratification: NR | Regular visits, allowing early discharge | Monitoring: face to face<br><br>Self-management: NR | Outcomes/aims: NR | CMOC10 (communication), CMOC11 (at home), CMOC12 (caregiver) |
| Rankin 2010 [35]<br><br>Wandsworth, England | Model 1a.<br>Urgency: not urgent<br>Duration in VW: NR | Focus on people with chronic conditions; but also patients >18 years of age and vulnerable to admission (includes drug & alcohol, | Initial (joint) assessment at patients home, Care plan, Patients | Monitoring: NR, but recommendation for future: integrate and | Outcomes/aims: To reduce emergency hospital admissions by supporting patients in the | CMOC4 (MDT meetings), CMOC8 (case management), CMOC10 (communication) |

| Study detail | Virtual Ward type | Population | Intervention | Monitoring, self-management | Outcomes/aims | CMOC addressed |
|---|---|---|---|---|---|---|
| Case study, mixed methods | Open 24/7? - not 24/7 (but future plans) | mental health – anyone)<br>Selection: risk tool for UHA; Patients at high risk of admission highlighted by PARR++ or combined risk tool (select patients >70% risk); GP referrals, Secondary Care (A&E, MAU, Geriatrics, Sickle Cell etc),<br>Intermediate Care Team, Community Nurses, Ambulance Services<br>Stratification: NR | discharged back to GP when considered no longer at risk of admission, Regular ongoing follow-up of patient at home. Weekly MDT meetings. Daily activity rounds with GP, community matrons & ward clerk (Co-ordinator) | expand Telehealth solutions<br>Self-management: NR | community. To pro-actively manage patients identified as being at risk of admission. To prevent patients being admitted and facilitate discharge: 'pull' patients out of hospitals rather than expect risk averse secondary care to 'push'; safe place to 'push' or 'pull' patients to | |
| Ryland 2015 [36]<br><br>Started Oct 2012 (study 2013)<br><br>Bradford, England<br><br>Case study | Model 2<br><br>Urgency: medical emergency – mainly step-down from hospital, but some step-up (according to Shepperd, having medical emergency).<br><br>Duration in VW: short-term (approx. 1 week – Shepperd)<br>Open 24/7? – implied 24h ('could get in touch 24h') | 41 patients per month (20 at any one time), with frailty<br><br>Selection: Frailty (no screening tool) + Acute change in health or functional status (mainly step-down) – Shepperd<br>Stratification: NR | Reactive? And Proactive? (CGA) | Monitoring: NR<br><br>Self-management: yes (enablement) | Outcomes/aims: hospital readmission, hospital length of stay, rate of admission into Geriatric Medicine beds, integration of services, recruitment/retention of community support team, pressure on acute hospital | CMOC2 (IT), CMOC11 (at home) |

| Study detail | Virtual Ward type | Population | Intervention | Monitoring, self-management | Outcomes/aims | CMOC addressed |
|---|---|---|---|---|---|---|
| Sheffield 2018 [37]<br><br>from 2017<br><br>Sheffield, England<br><br>Case study | Model 1a<br><br>Urgency: probably non-acute<br><br>Duration in VW: NR (implied longer term) NR<br><br>Open 24/7? – NR | Complex health conditions<br><br>Selection: data from Frailty index / Risk Stratification / Practice Staff intelligence (step-up) or secondary care email (step-down)<br><br>Stratification: Red/amber/green | Proactive.<br>Daily 'ward round' to monitor and action tasks (co-ordinator). Weekly MDT meeting. VW team and patient decides on VW admission and discharge. RAG rating reviewed by MDT. | Monitoring: NR<br><br>Self-management: NR | Outcomes/aims: provide wrap-around care to people in their own homes to reduce the need for hospital admission. | CMOC5 (Patient selection), CMOC10 (communication) |
| Swansea Bay 2020 [10]<br><br>started May 2020 (during COVID)<br><br>Swansea Bay, Wales<br><br>Case study | Model 1b<br><br>Urgency: before the situation reaches a crisis point; step up and step down.<br>Duration in VW: NR (implied not very short-term)<br><br>Open 24/7? - | Frailty, older people, and those with complex medical and social needs<br><br>Selection: Unclear, looks like clinical referral (patients with complex or multiple needs, who have a history of falls, frequent or recurrent hospital admissions, uncontrolled chronic conditions or health and social care needs)<br><br>Stratification: NR | Proactive (holistic, patient-centred, high quality care through rapid assessment; multidisciplinary team involvement and effective partnership working) | Monitoring: face to face; proactive (can be closely monitored to prevent accidents or deterioration)<br><br>Self-management: NR | Outcomes/aims: healthy living, stop deterioration, prevent admissions, earlier discharge | CMOC3 (MDT), |
